# Female-specific factors are associated with cognition in the UK Biobank cohort

**DOI:** 10.1101/2022.06.27.22276879

**Authors:** Linn R.S. Lindseth, Ann-Marie G. de Lange, Dennis van der Meer, Ingrid Agartz, Lars T. Westlye, Christian K. Tamnes, Claudia Barth

**Affiliations:** NORMENT, Institute of Clinical Medicine, University of Oslo, Oslo, Norway; LREN, Centre for Research in Neurosciences, Department of Clinical Neurosciences, Lausanne University Hospital (CHUV) and University of Lausanne, Lausanne, Switzerland; Department of Psychology, University of Oslo, Oslo, Norway; Department of Psychiatry, University of Oxford, Oxford, UK; NORMENT, Division of Mental Health and Addiction, Oslo University Hospital & Institute of Clinical Medicine, University of Oslo, Norway; School of Mental Health and Neuroscience, Faculty of Health, Medicine and Life Sciences, Maastricht University, the Netherlands; Department of Psychiatric Research, Diakonhjemmet Hospital, Oslo, Norway; Centre for Psychiatry Research, Department of Clinical Neuroscience, Karolinska Institutet & Stockholm Health Care Services, Stockholm County Council, Stockholm, Sweden; PROMENTA Research Center, Department of Psychology, University of Oslo, Oslo, Norway

**Author notes:** **Corresponding author:** Claudia Barth, PhD, E-Mail, postal address: Section Vinderen, Diakonhjemmet Hospital, P.O. Box 85, Vinderen, N-0319 Oslo, Norway.

## Abstract

**Background:** Relative to males, females are at a higher risk of developing age-related neurocognitive disorders including Alzheimer’s disease. Emerging evidence suggests that reproductive life events such as pregnancy and hormone use may influence female’s cognition later in life. Yet, female’s health has historically been understudied, and little is known about the relationship between female-specific factors and cognition.

**Methods:** Using multiple linear regression, we investigated the associations between reproductive history, exogenous hormone use, apolipoprotein (*APOE)* ε4 genotype and cognition in 221,124 middle-to older-aged (mean age 56.2 ± 8.0 years) females from the UK Biobank. Performance on six cognitive tasks was assessed, covering four cognitive domains: episodic visual memory, numeric working memory, processing speed, and executive function.

**Results:** A longer reproductive span, older age at menopause, older age at first and last birth, and use of hormonal contraceptives were positively associated with cognitive performance later in life. Number of live births, hysterectomy without oophorectomy and use of hormone therapy showed mixed findings, with task-specific positive and negative associations. Effect sizes were generally small (Cohen’s *d* < 0.1). While *APOE* ε4 genotype was associated with reduced processing speed and executive functioning, in a dose-dependent manner, it did not influence the observed associations between female-specific factors and cognition.

**Conclusion:** Our findings support previous evidence of associations between a broad range of female-specific factors and cognition. The positive association between a history of hormonal contraceptive use and cognition later in life showed the largest effect sizes (max. *d* = 0.1). Future research is needed to investigate the effects of sex hormone exposure and cognition to develop a better understanding of female’s brain health.

## Introduction

Numerous age-related neurocognitive disorders show prominent sex differences in their prevalence and presentation. For instance, relative to males, female are more likely to develop Alzheimer’s Disease (AD), and are afflicted with poorer cognitive outcomes ^1^. Female’s health, however, has historically been understudied, and little is known about the roles of female-specific factors, such as reproductive history and hormone use, for cognitive performance later in life ^2^.

Emerging epidemiological evidence suggests an association between reproductive span and cognitive function. A longer reproductive span (age at menarche until age at menopause) has been associated with better cognition later in life, such as better global cognition ^3-6^, verbal memory ^3^, executive function ^4^, and verbal fluency ^7^. However, other studies found no such associations ^7, 8^. Age at menarche ^4, 7^ and age at menopause ^9, 10^ have each been both negatively and positively associated with cognitive functioning. While a later natural transition to menopause (on average at 51.4 years^11^) may positively influence late life cognition, a recent systematic review suggests an association between surgical menopause, i.e., the removal of both ovaries (bilateral oophorectomy) before the onset of menopause, and faster cognitive decline in several cognitive domains ^12^. Similarly, a hysterectomy (removal of the uterus), often combined with bilateral oophorectomy, may also impact cognitive performance later in life ^12^. However, results are mixed.

Another factor which may alter both the length of the reproductive span^13^ and cognition is a history of childbirths. A higher number of childbirths may extend the length of a female’s reproductive span^14^, and has been linked to positive outcomes in brain imaging ^15-18^ and cognitive domains such as processing speed, visual memory ^19^, and verbal memory ^20^. However, other studies found no such associations ^4, 7^, or negative associations between parity and global cognition ^3, 6, 9^ and verbal memory ^3^. These discrepancies between findings may be linked to potential non-linear effects ^5, 19^. Compared to having two children, both no or one child and three or more children have been associated with poorer performance in verbal memory and executive function ^21^. Besides the number of childbirths, maternal age at first birth has also been associated with cognitive performance. For instance, older age at first birth has been linked to better cognition in previous studies ^7, 21^. However, this association may be influenced by socioeconomic factors ^22^.

In addition to the associations of reproductive span and parity with late-life cognition, hormone uses in the form of hormonal contraception (HC) to commonly prevent pregnancies or hormone therapy (HT) to alleviate menopausal symptoms may further modulate female’s late-life cognitive performance. Despite some studies to the contrary ^23^, the use of HT has largely been associated with protective effects on cognition across multiple domains, including conceptualization and visuo-practical skills ^24^, verbal memory ^25, 26^, visual memory ^7^, and global cognition ^6^. However, this positive association between HT use and cognition may be modulated by genotype, age at initiation and duration of use. Carried by 14% of the world’s population, the apolipoprotein E type 4 (*APOE* ε4) allele is a known risk factor for AD ^27^. Yaffe and colleagues found that current HT use lowered the risk of lower cognitive performance by almost half compared to never-users, but only in non-carriers ^28^. Similarly, results from the Nurses’ Health Study found that HT use was associated with faster decline in general cognition, especially among females with an *APOE* ε4 allele ^23^. Furthermore, according to the “*critical window hypothesis*”, the use of HT may be beneficial for cognition when initiated during perimenopause, while potentially detrimental if initiated later ^29-33^. However, other studies did not support this hypothesis ^23^. Whether duration of HT use modulates cognition beyond or independent of an individual’s age at initiation is unclear.

Relative to HT, the influence of HC on cognition is far less studied, despite its widespread use ^2^. Studies on HC use commonly contrast cognitive performance in pre-menopausal users and non-users at high or low hormone states across the menstrual cycle, linking HC use to better performance on overall cognition, immediate memory/learning, attention, delayed memory ^34^, and verbal memory ^35^. However, the results are inconclusive, largely due to small sample sizes with an overall mean of 24 HC users per study, including several studies with 10 or fewer participants ^36^. Only a limited number of studies have investigated the association between HC use and cognition later in life, again with mixed results: some report positive correlations between HC use and global cognition and verbal memory in late life ^4-6^, while others do not ^7, 9^. Data from the Wisconsin Registry for Alzheimer’s Prevention suggests a positive association between HC duration and cognitive performance ^37^. However, the sample of HC never-users was small (n = 34), precluding firm conclusions.

In summary, previous research indicates that reproductive history and hormone use may influence female’s cognition later in life. However, the findings have been inconclusive and both positive, negative and no associations have been reported, possibly due to a combination of small samples and various moderating factors. Here, we investigated the association of reproductive years, reproductive history, and HT and HC use with cognition in 221,124 middle-to older-aged females from the UK Biobank. Based on previous studies ^3, 4, 10, 19, 21^, we assumed that female-specific factor may be particularly associated with memory, executive function and processing speed. More specifically, we hypothesized that lower age at menarche, higher age at menopause and a longer reproductive span would be associated with better cognitive performance. Given past research on parity, we expected a non-linear relationship for number of live childbirths and cognition, with two live childbirths being associated with better cognitive scores compared to lower or higher numbers of live births. Concerning hormone use, we expected that HT and HC would be positively associated with cognition. Yet, the effects may be dependent on age at initiation and duration of use. To examine the effect of *APOE* ε4 genotype status, which has been suggested to modify the associations between female-specific factors and cognition, follow-up analyses including interaction terms were performed for each of the measures of interest.

## Methods and materials

### Participants

The sample was drawn from the UK Biobank cohort (https://www.ukbiobank.ac.uk), and included 273,384 females. Sex of participants refers to binary data on biological sex acquired at recruitment. To ascertain a cognitively healthy sample, participants with diagnosed brain disorders known to influence cognition were excluded from the main sample (n = 39,011, see supplementary Note 1), as were participants who later withdrew their consent (n = 6). Sample demographics of females with sufficient data on key demographic variables including age, education, ethnicity, Townsend deprivation index and lifestyle score, amounting to 221,124, are provided in Table 1. Sample demographics stratified by HT and HC user status are displayed in supplementary Table S1 and Table S2, respectively.

**Table 1.** Sample demographics.

|  |  |
| --- | --- |
| **Total N** | **221,124** |
| **Age** (years)* | 56.2 ± 8.0 |
| **Age range** (years) | 39 – 70 |
| **Education, N (%)** |  |
| College/University degree | 73,577 (33.5) |
| O levels/GCSEs or equivalent | 52,103 (23.7) |
| None of the above | 33,165 (15.1) |
| A levels/AS levels or equivalent | 27,167 (12.4) |
| Other professional qualifications | 12,764 (5.8) |
| CSEs or equivalent | 11,499 (5.2) |
| NVQ/HND/HNC or equivalent | 9,307 (4.2) |
| **Ethnicity, N (%)** |  |
| White | 210,498 (95.2) |
| Asian | 3,298 (1.5) |
| Black | 3,282 (1.5) |
| Chinese | 791 (0.4) |
| Other ethnic groups | 1,849 (0.8) |
| Mixed | 1,406 (0.6) |
| **Townsend Deprivation Score\*** | -1.5 ± 2.9 |
| **Lifestyle score\*** | 2.9 ± 1.6 |

\*Continuous data in mean ± standard deviation and categorical data as number (%). Abbreviations: N = Number; O = Ordinary Level Qualification; GCSE = General Certificate of Secondary Education; A = Advanced Level Qualification; AS = Advanced Subsidiary Level Qualification; CSE = Certificate of Secondary Education; NVQ = National Vocational Qualification; HND = Higher National Diploma; HNC = Higher National Certificate

### Cognitive assessment

Six computerized cognitive tests that were completed by more than 10% of the whole study population at baseline were selected. The included tests covered 4 cognitive domains, namely *visual memory* (Pairs Matching Test, N = 220,084, number of errors), *working memory* (Digit Span Test, N = 22,184, number of correct digits), *processing speed* (simple - Reaction Time Test, N = 220,098, in milliseconds; complex - Symbol Substitution Test, N = 58,083, number of correct substitutions), and *executive function* (Trail Making Test A & B, N = 50,464, in deciseconds). The cognitive tests are described in detail elsewhere ^38, 39^. All test scores, except for the symbol digit test, were log-transformed. As the raw scores for the pair matching test and digit span test included zero values, plus one was added before transformation. For reaction time, potential outliers with scores below 50 milliseconds and above 2000 milliseconds were removed. For the Symbol Digit Test, scores below three and above 36 were removed as outliers. To ease interpretation of results, Reaction Time, Trail Making A and B, as well as Pair Matching scores were inverted (multiplied by -1) so that higher scores on all the cognitive tests reflect better performance.

### Assessment of female-specific factors

Female-specific variables included length of reproductive span in years (age at menopause - age at menarche), age at menopause, age at menarche, number of live births, age at first and last childbirth, history of pregnancy loss during 1^st^ trimester (miscarriage/termination) and after the 2^nd^ trimester (stillbirth), history of and age at bilateral oophorectomy and/or hysterectomy, HT and HC usage status (current-user/past-user/never-user), and duration of use and age at initiation among the users. Participants who had missing data, or had responded “do not know”, “prefer not to answer”, “none of the above” or similar for each of the relevant variables, were excluded for the respective analyses.

### Genotyping

To assess *APOE* genotype, we used the extensively quality-controlled UK Biobank version 3 imputed data ^40^. *APOE* ε genotype was approximated based on two *APOE* ε single□nucleotide polymorphisms—rs7412 and rs429358, in accordance with previous work ^41^. APOE ε4 status was labeled *carrier* for ε3/ε4 and ε4/ε4 combinations, and *non□carrier* for ε2/ε2, ε2/ ε3, and ε3/ ε3 combinations ^42^. To test for potential dose-dependent effects, ε3/ε4 was labeled carrier of one ε4 allele, and ε4/ε4 as carrier of two ε4 alleles. Due to its ambiguity with ε1/ε3, the homozygous ε2/ε4 allele combination was removed ^43^. Sample demographics stratified by *APOE* ε4 status are displayed in supplementary Table S3.

### Statistical analysis

Multiple linear regression analyses were performed to investigate the relationship between female-specific variables (independent variable) and cognitive test scores (dependent variable).

All models included the following additional independent variables known to influence reproductive history, hormone use and cognition: age, education, body mass index (BMI), Townsend deprivation index, and lifestyle score ^44-46^ (see supplementary Note 2 for details on Townsend deprivation index and lifestyle score). In addition, the analyses for reproductive span and age at menopause were corrected for use of HT, use of HC, history of bilateral oophorectomy and/or hysterectomy, and number of live births.

We also investigated potential non-linear effects of the number of childbirths on cognition, by adding a quadratic term to the model, and by adding number of childbirths (0, 1, 2, 3, 4, 5, 6, 7-22) as a dummy variable instead of a continuous variable. In latter model, participants with zero childbirths served as a reference group. In addition, we tested whether age at first childbirth and age at last childbirth were associated with cognitive performance in parous individuals. To further examine whether having been pregnant, without term birth and parental experience, might influence cognition, we also contrasted individuals who had zero live births, but one or more miscarriages and terminations during the first trimester or stillbirths (i.e., baby is born dead after 24 weeks of pregnancy) after the 2^nd^ trimester, against individuals who never were pregnant.

To examine whether cognitive performance was associated with a history of bilateral oophorectomy, as a proxy of surgical menopause, as well as hysterectomy without bilateral oophorectomy, we fitted additional linear regression models adjusting for basic covariates and HT user status. We further interrogated whether age at oophorectomy/hysterectomy influenced cognition later in life.

Lastly, additional multiple linear regression models were fitted including an *APOE* ε4 status□ ×female-specific measure interaction term to assess the associations between *APOE* ε4 status and female-specific factors on cognition. The models were adjusted for the same covariates as listed above. Furthermore, we tested for main effects of APOE ε4 status on cognitive performance, again adjusting for the same covariates.

The statistical analyses were conducted using R, version 4.1.2 ^47^. All variables were standardized (subtracting the mean and dividing by the SD) prior to the regression analyses. To account for multiple comparisons, the p-values are reported before and after correcting for false discovery rate (p_FDR_, across all main models). The significance threshold was set to α = 0.05. We computed the Cohen’s *d* effect sizes from the *t*-statistics for categorical variables and via the partial correlation coefficient (*r*) for continuous variables ^48^. Correlations between demographics, female-specific factors and cognitive tests are shown in Figure 1.

**Figure 1.**
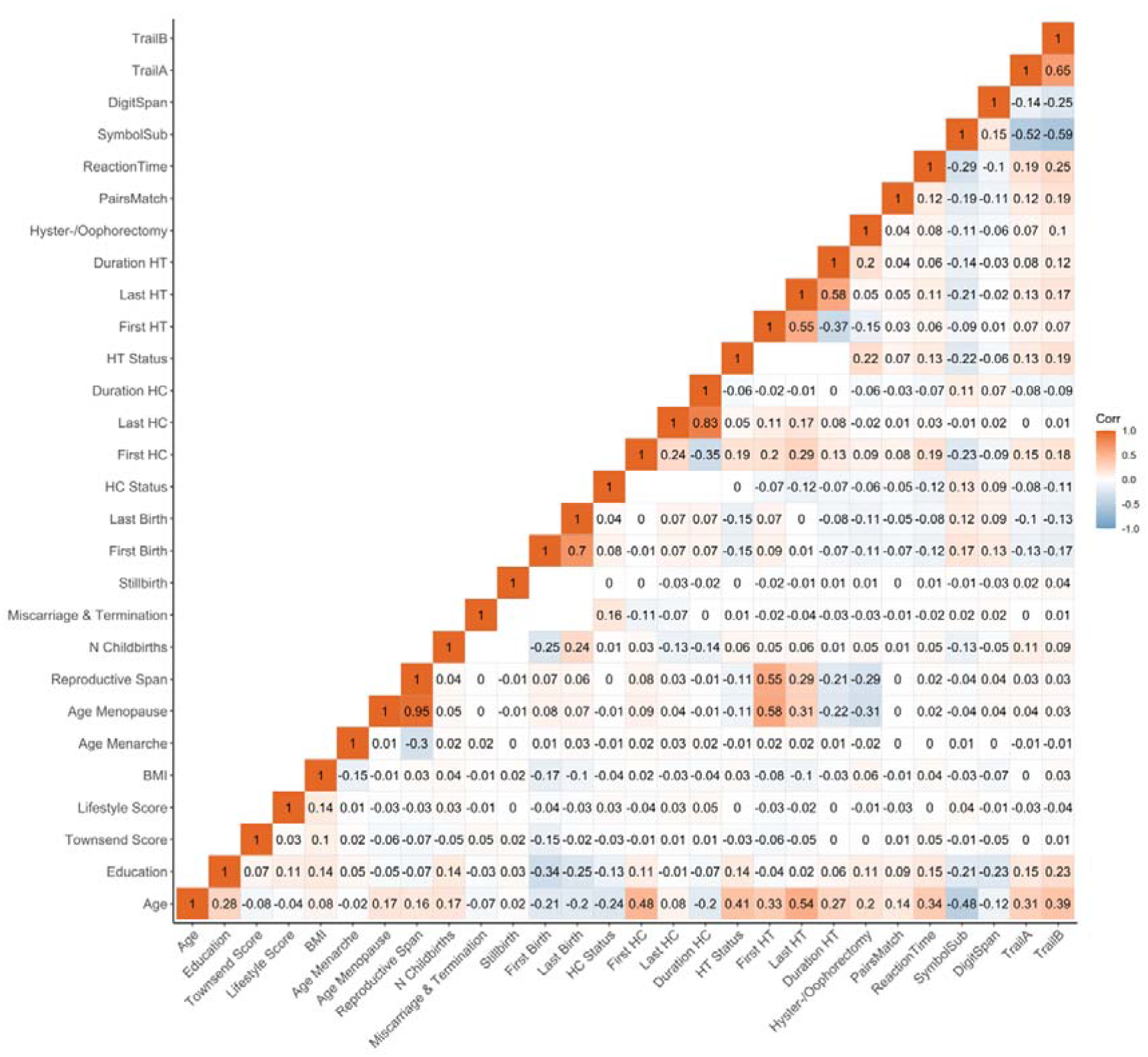
Correlations (Pearson’s r) between demographics, female-specific factors and cognitive test scores. Correlation between each pair of variables is computed using all complete pairs of observations on those variables. Empty fields indicate no complete pairs for that pair of variables. Abbreviation: BMI = body mass index, HT = hormone therapy, HC = hormonal contraceptive, N = Number, First Birth = Age at first live birth (years), Last Birth = Age at last live birth (years), First HC = Age at first HC use (years), Last HC = Age at last HC use (years), First HT = Age at first HT use (years), Last HT = Age at last HT use (years).

### Sensitivity analyses

To control for the potential influence of extreme values on our results, we assessed each continuous female-specific factor for outliers and excluded the corresponding participants before re-running the respective main analysis. To identify extreme values, we applied the median absolute deviation (MAD) method, implemented in the R package *Routliers* (https://CRAN.R-project.org/package=Routliers), using default settings (i.e., MAD threshold of ± 3). This approach has the advantage of being robust with respect to sample size and the presence of extreme values. Furthermore, the main models were re-run (1) including previously excluded participants with diagnosed brain disorders to test whether results are sensitive to potential selection biases, and (2) also adjusting for *APOE* ε4 status.

## Results

Significant associations between female-specific factors and cognitive performance are displayed in Figure 2. All tested associations are visualized in supplementary materials, Figure S1, and Table S4.

**Figure 2.**
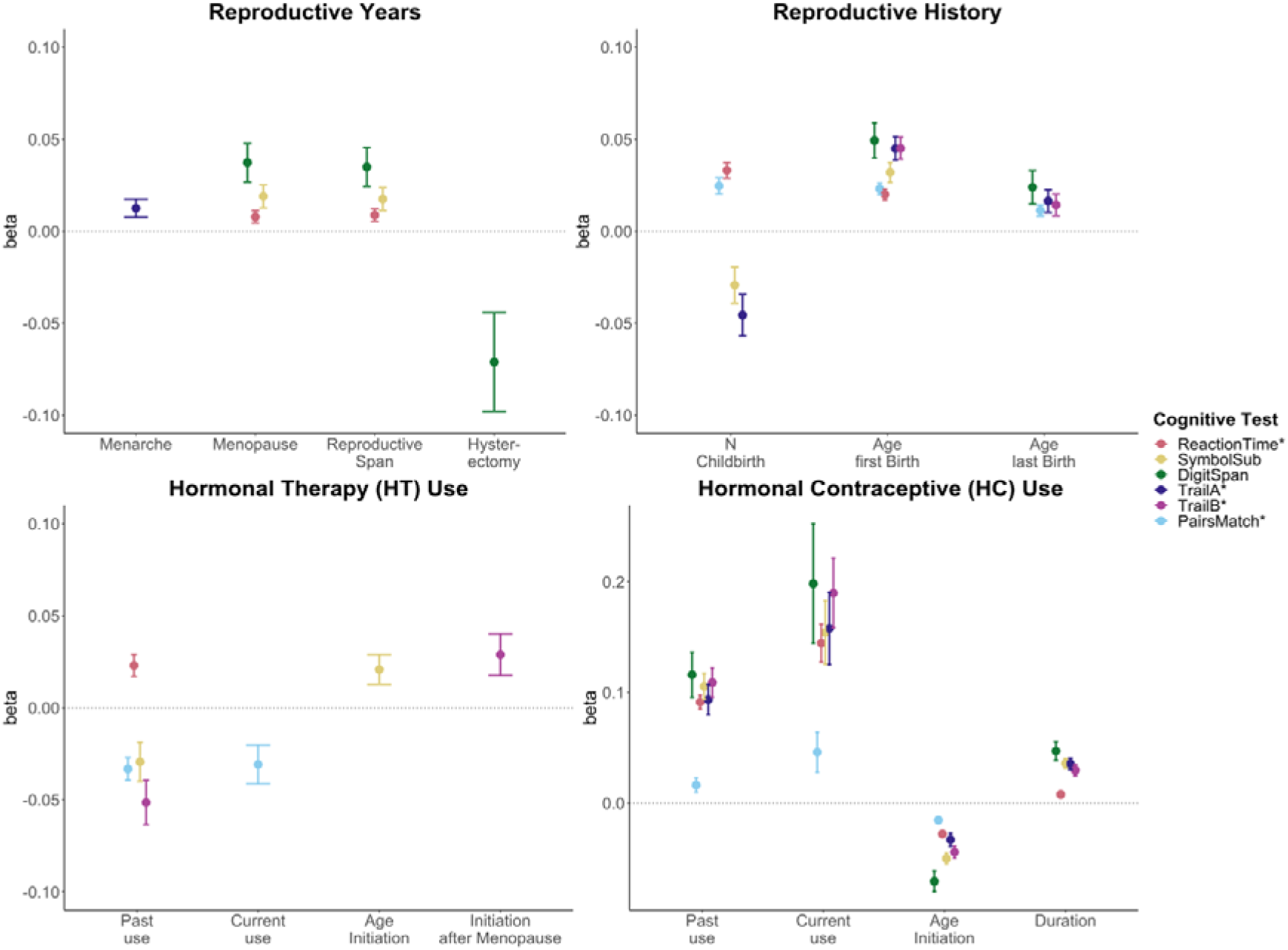
Significant associations between female-specific factors and cognitive performance. Point plot of beta-values with standard error from separate multiple regression analysis with cognitive task as dependent variable and female-specific variables as independent variable. All models are adjusted for age, education, body mass index, Townsend deprivation score, lifestyle score. In addition, the analyses for reproductive span and age at menopause were corrected for use of HT, use of HC, history of hysterectomy and bilateral oophorectomy, and number of live births. The HT models were additionally adjusted for history of hysterectomy and bilateral oophorectomy, and the hysterectomy/oophorectomy model was co-varied for use of HT. All variables were standardized prior to performing the multiple linear regression analysis (subtracting the mean and dividing by the standard deviation). For visualization purposes, cognitive tests marked with * are inverted (multiplied by -1) so that positive beta-values always indicate associations between higher values on the female-specific variables and better performance on cognitive tests.

### Reproductive years and cognitive performance

A longer *reproductive span* was associated with faster processing speed (Reaction Time: *β* = 0.009, p = 0.009, p_FDR_= 0.025, Cohen’s *d* = 0.017; Symbol Substitution: *β* = 0.017, p = 0.005, p_FDR_= 0.016, *d* = 0.035), and higher working memory scores (Digit Span: *β* = 0.035, p = 0.001, p_FDR_= 0.003, *d* = 0.066). Similar positive associations were found for *age at menopause*. However, older *age at menarche* was associated with higher executive function scores (Trail Making A: *β* = 0.013, p = 0.010, p_FDR_= 0.028, *d* = 0.025).

While a history of hysterectomy, without bilateral oophorectomy, was associated with lower working memory scores (Digit Span: *β* = -0.071, p = 0.008, p_FDR_= 0.025, *d* = -0.071), bilateral oophorectomy, as a proxy of surgical menopause, was not significantly associated with cognitive performance, after FDR correction. Similarly, after correction, *age at bilateral oophorectomy* and/or *age at hysterectomy* were not significantly associated with cognitive scores.

### Reproductive history and cognitive performance

A higher *number of live childbirths* was associated with faster simple processing speed (Reaction Time: *β* = 0.033, p = 6.99e-15, p_FDR_= 1.03e-13, *d* = 0.037), and higher visual memory scores (Pair Matching: *β* = 0.025, p = 1.82e-08, p_FDR_ = 1.05e-07, *d* = 0.027). However, a higher number of live childbirths was also associated with slower complex processing speed (Symbol Substitution: *β* = -0.029, p = 0.003, p_FDR_ = 0.010, *d* = -0.027) and lower executive functioning scores (Trail Making A: *β* = -0.046, p = 6.29e-05, p_FDR_ = 2.68e-04, *d* = -0.039).

For visual memory and simple processing speed, we found a significant non-linear association with the number of live childbirths. Follow-up multiple linear regression models including number of live childbirths as a categorical variable showed that up to three and four childbirths were associated with faster simple processing speed and higher visual memory scores, respectively. More than four childbirths were associated with slower simple processing speed (Figure 3, supplementary Table S5). In parous individuals, an older *age at first childbirth* was associated with higher cognitive performance scores on all six tests. Similarly, older *age at last childbirth* was associated with higher scores for visual memory, working memory and executive functioning.

**Figure 3.**
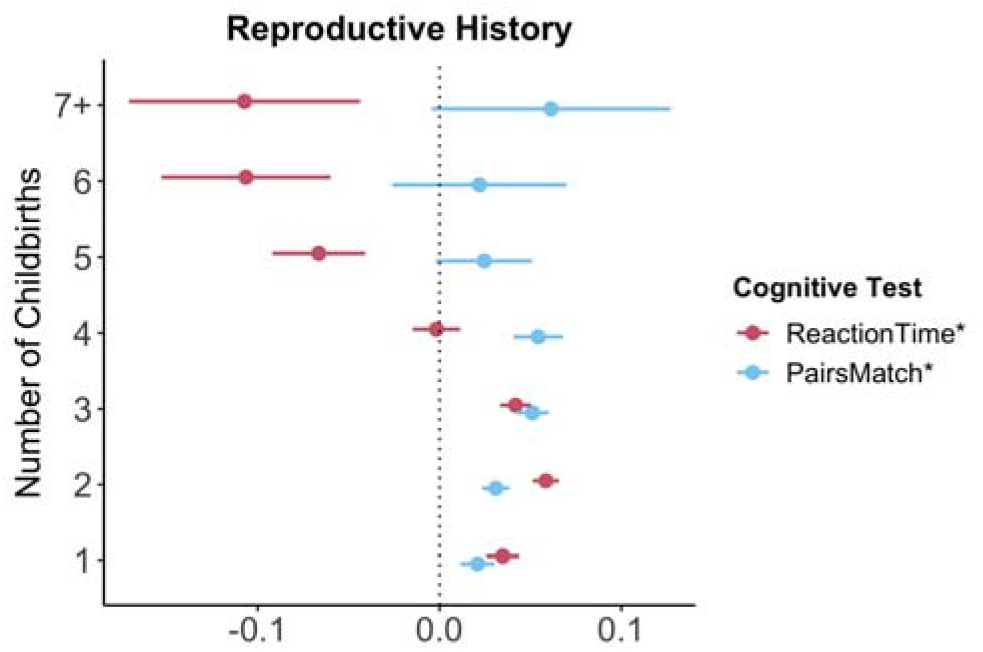
Significant non-linear association between number of childbirths and cognitive performance. Point plot of beta values with standard error from multiple regression analyses with cognitive task as dependent variable and number of childbirths as categorical independent variable. All models are adjusted for age, education, body mass index, Townsend deprivation score, lifestyle score. All variables were standardized prior to performing the multiple linear regression analysis (subtracting the mean and dividing by the standard deviation). For visualization purposes, cognitive tests marked with * are inverted (multiplied by -1) so that positive beta-values always indicate associations between higher values on the female-specific variables and better performance on cognitive tests. Sample size for Pair Matching (0 = 34,881, 1 = 23,387, 2 = 77,687, 3 = 30,478, 4 = 7,596, 5 = 1,569, 6 = 446, 7+ = 234). Sample size for Reaction Time (0 = 34,699, 1 = 23,232, 2 = 77,308, 3 = 30,262, 4 = 7,520, 5 = 1,545, 6 = 430, 7+ = 226).

Relative to nulliparous individuals, a *history of pregnancy loss* during 1^st^ (miscarriage/termination) and after the 2^nd^ trimester (stillbirth), without a history of live births, was not associated with cognitive performance, after FDR-correction.

### HT and cognitive performance

*Past HT use* was associated with faster simple processing speed (Reaction Time: *β* = 0.023, p = 9.19e-05, p_FDR_= 3.79e-04, *d* = 0.036), but slower complex processing speed (Symbol Substitution: *β* = -0.029, p = 0.006, p_FDR_ = 0.018, *d* = -0.046). Past HT use was also associated with lower executive functioning scores (Trail Making B: *β* = -0.051, p = 1.72e-05, p_FDR_ = 7.56e-05, *d* = -0.048). Both *past* and *current HT use* were associated with lower visual memory scores (Pair Matching; *past use*: *β* = -0.033, p = 6.89e-08, p_FDR_ = 3.64e-07, *d* = -0.048; *current use*: *β* = -0.031, p = 0.004, p_FDR_ = 0.012, *d* = -0.026). While *duration of HT use* was not significantly associated with any of cognitive tests, an older *age at HT initiation* was associated with faster complex processing speed (Symbol Substitution: *β* = 0.021, p = 0.010, p_FDR_ = 0.027, *d* = 0.042). We further tested whether *age at HT initiation relative to age at menopause* (age started HT – age at menopause) was associated with late life cognition. *HT initiation after the onset of menopause* was associated with higher executive function scores (Trail Making B: *β* = 0.029, p = 0.010, p_FDR_ = 0.028, *d* = 0.054).

### HC and cognitive performance

*Current and past use of HC* were significantly associated with higher scores on all six cognitive tests. A longer *duration of HC use* was significantly associated with higher performance score in five of the six cognitive tests, except visual memory. An *older age at HC initiation* was associated with lower performance scores in all cognitive tests assessed.

#### Sensitivity analyses

Most results were robust after either (1) removing extreme values (Supplementary Table S6), (2) including previously excluded participants with known brain disorders (Supplementary Table S7), and (3) additionally adjusting for *APOE* ε4 status (Supplementary Table S8), with slight variations. Detected extreme values are highlighted in Supplementary Table S9.

### *APOE* ε4 genotype and cognitive performance

Relative to non-carriers, carriers of *APOE* ε4 alleles showed significantly lower executive functioning scores (Trail B: *β* = -0.028, p = 0.008, p_FDR_ = 0.027, *d* = -0.030) and slower complex processing speed (Symbol substitution: *β* = -0.046, p = 6.59e-07, p_FDR_ = 5.93e-06, *d* = -0.053).

Follow-up analyses suggest dose-dependent effects of *APOE* ε4 genotype on executive functioning (Trail B: two ε4 alleles *β* = -0.111, p = 2.96e-04, p_FDR_ = 0.001, *d* = -0.107; one ε4 allele *β* = -0.020, p = 0.064, p_FDR_ = 0.123, *d* = -0.055), with carriers of two ε4 alleles, not one allele, showing lower cognitive performance scores (see Figure 4). Relative to non-carriers, slower complex processing speed was found in carriers with one and two ε4 alleles (Symbol substitution: two ε4 alleles *β* = -0.145, p = 1.33e-07, p_FDR_ = 2.39e-06, *d* = -0.057; one ε4 allele *β* = -0.037, p = 1.23e-04, p_FDR_ = 7.39e-04, *d* = -0.042, see Supplementary Table S10).

**Figure 4.**
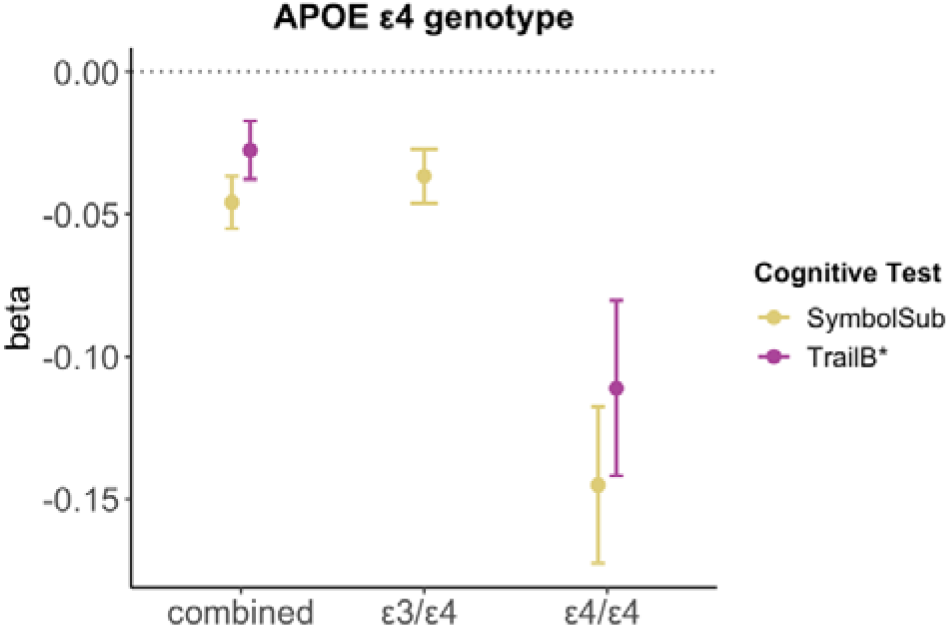
Significant associations between *APOE* ε4 genotype and cognitive performance. Point plot of beta-values with standard error from multiple regression analyses with cognitive task as dependent variable and APOE e4 genotype as categorical independent variable. All models are adjusted for age, education, body mass index, Townsend deprivation score, lifestyle score. All variables were standardized prior to performing the multiple linear regression analysis (subtracting the mean and dividing by the standard deviation). For visualization purposes, cognitive tests marked with * are inverted (multiplied by - 1). Sample size for Symbol Substitution (non-carrier = 34,266, carrier one e4 allele = 11,023, carrier two e4 allele = 1,028) and Trail Making B (non-carrier = 29,865, carrier one e4 allele = 9,559, carrier two e4 allele = 893).

We found no significant interactions between *APOE* ε4 genotype and female-specific factors on cognition (Supplementary Table S11), except for number of live childbirths as well as number of live childbirths squared and simple processing speed (Reaction time: N Childbirths *β* = 0.039, p = 3.32e-04, p_FDR_ = 0.022, N Childbirths^2^ *β* = -0.046, p = 1.28e-04, p_FDR_ = 0.017). However, after excluding extreme value (number of childbirths > 6), the interaction did not survive correction for multiple comparison.

## Discussion

The results of the present study indicate that a longer reproductive span, older age at menopause, older age at first and last childbirth, and use of hormonal contraceptives (HC) are positively associated with cognition later in life. Number of live births, hysterectomy without oophorectomy, and use of hormone therapy showed mixed findings, with task-specific positive and negative associations. Most of the results remained significant after additional sensitivity analyses. While effect sizes were generally small (*d* < 0.1), the association between a history of HC use and cognition later in life showed the largest effect sizes (max. *d* = 0.1).

A longer reproductive span with an older age at menopause was associated with better simple processing speed, complex processing speed, and numeric working memory. Older age at menarche was also associated with faster complex processing speed later in life. Our results align with a number of previous studies suggesting a positive association between reproductive span and cognition ^3-7^. Studies on the effect of age at menarche on late-life cognition are inconclusive ^4, 6, 7^, and may be confounded by an inaccurate recall of age at menarche ^49^, especially in middle-to older-aged populations. Several previous studies found a positive association between higher age at menopause and cognition ^6, 9, 10^. Yet, it should be noted that the causal direction for this association is unclear. One study found a positive association between cognitive scores in childhood and age at menopause, suggesting that cognitive function in childhood may influence age at menopause via early environmental or genetic programming ^50^. The present study did not have data on childhood cognition.

A history of hysterectomy, without bilateral oophorectomy, was associated with lower working memory scores. Bilateral oophorectomy, as proxy of surgical menopause, and age at hysterectomy and/or oophorectomy were not associated with cognitive performance later in life. While some studies reported no significant associations between cognition and proxies of surgical menopause ^7-9^, a recent systematic review associated surgical menopause at any age with a faster decline in verbal and semantic memory, and processing speed ^51^. However, the definition of surgical menopause varied across studies, rendering firm conclusions difficult. In general, our findings for reproductive years align with another study using data from UK Biobank, showing that a shorter reproductive span, younger age at menopause and a history of hysterectomy were associated with greater risk for all-cause dementia ^52^. Similarly, a shorter reproductive span with early age at natural or surgical menopause has also been linked to a higher risk of incident cardiovascular disease ^53^.

The observed positive association between longer reproductive span and better late-life cognition might be understood through the protective role of estradiol, the most prevalent and potent estrogen in the female body ^54^. Menopause is characterized by a cessation of ovarian function and a subsequent estradiol withdrawal. One might speculate that a longer reproductive span with an older age at menopause results in a higher life-time exposure to estradiol, which has been linked to positive effects on cognition ^55^. Yet, female’s estradiol exposure is modulated, among others, by their reproductive history ^2, 56^, and the impact of parity on cognition is inconclusive.

We found that a higher number of live births was associated with both *better* visual memory scores and faster simple processing speed, and *slower* complex processing speed and lower executive functioning scores. In addition, we showed a non-linear relationship between number of live childbirths and simple processing speed as well as visual memory: having two to three live births was associated with better late life cognition than having one and more than four. The positive effects of parity on visual memory and simple processing speed corroborate previous results ^19, 20^. While a number of studies suggest a non-linear effect of number of childbirths on cognition ^5, 21^ and all-cause dementia ^52^, other studies did not replicate these associations ^19^. Further studies are needed to confirm if any positive effect of parity on late-life cognition may be less pronounced in single-and grand-parous individuals.

Older age at first and last childbirth was associated with better cognitive functioning in multiple domains. While these findings are in line with several studies ^7, 21^, previous reports have been mixed ^4, 6^. In the current study, older age at first and last birth was correlated with higher education levels (see Figure 1). This is in line with previous research highlighting that older motherhood has been linked to higher socioeconomic status, total income, educational attainment, labor force participation and lower total number of childbirths^22, 57^. Hence, although we account for socioeconomic factors and education in our models, our results may be influenced by the great number of psychosocial variables associated with older motherhood and cognition.

A meta-analysis on the impact of pregnancy on memory function found that both pregnant and postpartum individuals showed lower scores in memory measures that place high demands on executive cognitive control ^58^. Yet, it is unclear how long these domain-specific cognitive disturbances persist. Here, we found that complex processing speed and executive functioning scores were lower in middle-to-older aged females with a history of live births, suggesting that these cognitive functions may be altered long after the last birth.

The differential relationship between parity and cognition might be understood through a number of potential physiological and psychosocial mechanisms ^56^. First, pregnancy-related variations in estradiol might exert effects on cognition. Estradiol increases 300-fold during pregnancy, drops rapidly postnatally, and remains lower in parous individuals compared to nulliparous individuals all through menopause ^56^. Higher levels of estradiol have been linked to better visual memory ^59^. Hence, lower estradiol levels in parous individuals might explain the observed negative association between parity and cognition. Second, the *“pregnancy compensation hypothesis”* suggests that a history of childbirths has a dampening effect on the otherwise ramped up female immune system throughout adulthood^45^. In this view, if pregnancies fail to occur, the immune system becomes overly activated and starts releasing auto-antibodies that attack healthy cells ^60^, giving rise to autoimmune diseases and AD. In line with immunologic effects, more time spent pregnant in the first trimester has been associated with lower risk for AD ^61^. This may explain the apparent positive effects of parity on cognition. However, we did not find an association between pregnancy loss during the first and after the second trimester and late life cognitive performance.

The relationship between parity and cognition might also be understood through maternal behavior adaptation. For instance, pregnancy and postpartum have been associated with heightened neuroplasticity to facilitate maternal behavior and caring for the offspring ^56, 62^. For instance, better simple processing speed and visual memory in parous individuals may be behavioral adaptations to quickly respond to the needs of the child and to detect potential threads through enhanced associative memory encoding and retrieval, respectively. However, one might also speculate that being on alert to protect the child will reduce the capacity for mental flexibility, a part of executive function ^63^. Growing evidence suggests heightened plasticity in the maternal brain, increasing responsiveness to both negative and positive experiences ^64^. Hence, factors modulating maternal experiences such as environment, genetics, and personality traits, may be drivers for how the maternal brain changes.

Both current and past use of HT were associated with lower visual memory scores. Past use of HT was also associated with higher simple processing speed, but lower complex processing speed and executive functioning scores. Duration of HT use did not influence late life cognition. However, older age at HT initiation was associated with faster complex processing speed. While a large number of previous studies reported on positive effects of HT use on cognition ^6, 7, 26, 28^, a review covering the same six cognitive tasks of the present study also found both positive and negative associations with HT use ^26^. The heterogeneity in results is likely driven by different study designs (e.g., observational vs. experimental), sample sizes and adjusted covariates ^26^. In addition, detailed information on HT formulation, modes of administration (oral, transdermal or vaginal) and dosage is rarely available; all factors which might modulate HT effects on cognition ^32, 65^. Furthermore, timing of HT initiation, *APOE* ε4 genotype and socioeconomic as well as lifestyle factors may also modulate the HT effect on late life cognition. We found higher complex processing speed with older age at HT initiation and higher executive functioning scores with HT initiation after menopause, which is not in line with the “*critical window hypothesis*”, stating that HT is most beneficial for cognitive performance when administered perimenopausal^66^. Age at HT initiation is intrinsically linked to age at menopause (r = 0.58, Figure 1), as HT is prescribed to alleviate menopausal symptoms or replenish endogenous sex hormone levels after bilateral oophorectomy and/or hysterectomy. Our results suggest that both an older age at menopause and an older age at first HT use may be positively associated with complex processing speed later in life.

While we found a slower complex processing speed and lower executive functioning scores in *APOE* ε4 carriers relative to non-carriers, in line with previous work ^43^, we found no significant interaction effect between *APOE* ε4 genotype and HT use on cognition. The latter result is in line with some studies ^30^. Yet others found less cognitive decline amongst HT using non-carriers ^23, 28^. As the cognitive tests varied between studies, one might speculate that the previously observed HT-genotype interaction may be domain-specific. Levels of socioeconomic deprivation, measured with the Townsend deprivation index, and lifestyle score differed between HT users and non-users (see Table 2). According to the *healthy user bias*, individuals using HT tend to be healthier and better educated, particularly in observational studies ^67, 68^. This was not the case in the current study, potentially explaining test-specific negative associations with HT use, even after confound correction.

Contrary to the HT findings, we found that current and past use of HC were associated with higher performance on all cognitive tasks assessed. Younger age at HC initiation and a longer duration of use were significantly associated with higher cognitive performance scores later in life. While most studies in premenopausal females commonly report on no or inconclusive associations between HC use and a variety of cognitive tasks ^36^, two studies in menopausal females found duration-dependent increases in cognitive performance in HC users compared to never-users, especially in users with over 10 or 15 years of use ^4, 37^. Our results corroborate these findings in middle-to older-aged individuals. Emerging evidence suggests that different synthetic HC analogues affect cognition differently ^36^, which highlights the importance of accounting for HC formulations. This is lacking in the present study. The mechanisms behind the positive associations with long-term HC and cognition are far from understood and warrant further exploration, especially as many females start using HCs at a young age and continue using it for decades.

To the best of our knowledge, the current work is one of the largest, comprehensive studies of the associations between female-specific factors, *APOE* ε4 genotype, and cognition. Large-scale population-based studies enable the identification of subtle effects that could go undetected in smaller samples, and are key to identify factors that may contribute to cognitive aging and risk for neurodegenerative disease. However, the cross-sectional and non-experimental nature of the presented data does not enable causal inference, and randomized controlled longitudinal studies are required to fully understand how female-specific factors influence female’s late life cognition. Furthermore, the cognitive test battery was limited, tapping only selected cognitive domains and functions. More research with a broader and more fine-grained array of cognitive tests is needed to elucidate the effects of female-specific factors on female’s cognitive functioning. As highlighted above, the current study lacked details on HT and HC formulation, mode of delivery (e.g., oral or transdermal), and dosage. Of note, HT and HC formulations and dosages have changed significantly over time. For instance, while present day HCs typically contain 15 to 35 μg of ethinyl estradiol and lower amounts of progestin, estrogen content was up to ∼150 μg and 1 to 10 mg of progestin when HCs were first introduced in the 1960s ^69^. Given the age range of the UK Biobank cohort at baseline, 40 – 70 years, considerable differences in HC and HT dosages by age when first used may be present, and results from the current study may not translate to other cohorts.

Furthermore, while we account for socioeconomic and lifestyle factors known to influence reproductive history, hormone use and cognition, it is possible that other factors such as childhood cognition, nutrition and physical/somatic health also influence the association between female-specific factors and late-life cognition^50^. In addition, the present work relies on self-reported data for hormone use and reproductive events, which might not always be reliable.

In summary, this study provides evidence for associations between female-specific factors and late life cognition. Specifically, longer reproductive span, higher age at menopause, higher age at first and last birth, and use of HC were associated with better cognitive performance later in life. The results for number of live births, hysterectomy and/or bilateral oophorectomy and use HT were mixed, with both positive and negative associations.

Future research is needed to investigate the effects of sex hormone exposure and cognition to develop a better understanding of female’s brain health.

## Data Availability

The data that support the findings of this study are available through the UK Biobank data access procedures.

https://www.ukbiobank.ac.uk/researchers

## Notes

### Competing Interest Statement

The authors have declared no competing interest.

### Funding Statement

While working on this study, the authors received funding from the Research Council of Norway (Christian K. Tamnes: #223273, #288083, #323951; Lars T. Westlye: #273345, #249795, #298646, #300768, #223273; Ingrid Agartz: #213700, #223273, #250358), the South-Eastern Norway Regional Health Authority (Christian K. Tamnes: #2019069, #2021070, #500189; Lars T. Westlye: #2018076, #2019101, Ingrid Agartz: #2017097, #2019104, #2020020), the European Research Council under the European Union's Horizon 2020 research and innovation programme (Lars T. Westlye: #802998), the Swiss National Science Foundation (Ann-Marie G. de Lange: PZ00P3_193658).

### Author Declarations

The data that support the findings of this study are available through the UK Biobank data access procedures (https://www.ukbiobank.ac.uk/researchers)

## References

1. Laws KR, Irvine K, Gale TM. Sex differences in Alzheimer’s disease. Curr Opin Psychiatry 2018; 31(2): 133–139.

2. Taylor CM, Pritschet L, Yu S, Jacobs EG. Applying a Women’s Health Lens to the Study of the Aging Brain. Front Hum Neurosci 2019; 13: 224.

3. Heys M, Jiang C, Cheng KK, Zhang W, Au Yeung SL, Lam TH et al. Life long endogenous estrogen exposure and later adulthood cognitive function in a population of naturally postmenopausal women from Southern China: the Guangzhou Biobank Cohort Study. Psychoneuroendocrino 2011; 36(6): 864–873.

4. Karim R, Dang H, Henderson VW, Hodis HN, St John J, Brinton RD et al. Effect of Reproductive History and Exogenous Hormone Use on Cognitive Function in Mid- and Late Life. J Am Geriatr Soc 2016; 64(12): 2448–2456.

5. Li FD, He F, Chen TR, Xiao YY, Lin ST, Shen W et al. Reproductive history and risk of cognitive impairment in elderly women: a cross-sectional study in eastern China. J Alzheimers Dis 2016; 49(1): 139–147.

6. Song X, Wu J, Zhou Y, Feng L, Yuan JM, Pan A et al. Reproductive and hormonal factors and risk of cognitive impairment among Singapore Chinese women. Am J Obstet Gynecol 2020; 223(3): 410 e411-410 e423.

7. Ryan J, Carriere I, Scali J, Ritchie K, Ancelin ML. Life-time estrogen exposure and cognitive functioning in later life. Psychoneuroendocrino 2009; 34(2): 287–298.

8. Low LF, Anstey KJ, Jorm AF, Rodgers B, Christensen H. Reproductive period and cognitive function in a representative sample of naturally postmenopausal women aged 60-64 years. Climacteric 2005; 8(4): 380–389.

9. McLay RN, Maki PM, Lyketsos CG. Nulliparity and late menopause are associated with decreased cognitive decline. J Neuropsychiatry Clin Neurosci 2003; 15(2): 161–167.

10. Kuh D, Cooper R, Moore A, Richards M, Hardy R. Age at menopause and lifetime cognition: Findings from a British birth cohort study. Neurology 2018; 90(19): e1673–e1681.

11. Brinton RD, Yao J, Yin F, Mack WJ, Cadenas E. Perimenopause as a neurological transition state. Nat Rev Endocrinol 2015; 11(7): 393–405.

12. Puri BK, Hutton SB, Saeed N, Oatridge A, Hajnal JV, Duncan LJ et al. A serial longitudinal quantitative MRI study of cerebral changes in first-episode schizophrenia using image segmentation and subvoxel registration. Psychiat Res-Neuroim 2001; 106(2): 141–150.

13. Mishra GD, Pandeya N, Dobson AJ, Chung HF, Anderson D, Kuh D et al. Early menarche, nulliparity and the risk for premature and early natural menopause. Hum Reprod 2017; 32(3): 679–686.

14. Dorjgochoo T, Kallianpur A, Gao YT, Cai H, Yang G, Li H et al. Dietary and lifestyle predictors of age at natural menopause and reproductive span in the Shanghai Women’s Health Study. Menopause 2008; 15(5): 924–933.

15. Voldsbekk I, Barth C, Maximov, II, Kaufmann T, Beck D, Richard G et al. A history of previous childbirths is linked to women’s white matter brain age in midlife and older age. Hum Brain Mapp 2021; 42(13): 4372–4386.

16. de Lange AG, Kaufmann T, van der Meer D, Maglanoc LA, Alnaes D, Moberget T et al. Population-based neuroimaging reveals traces of childbirth in the maternal brain. Proc Natl Acad Sci U S A 2019; 116(44): 22341–22346.

17. de Lange AG, Barth C, Kaufmann T, Maximov, II, van der Meer D, Agartz I et al. Women’s brain aging: Effects of sex-hormone exposure, pregnancies, and genetic risk for Alzheimer’s disease. Hum Brain Mapp 2020.

18. de Lange AG, Barth C, Kaufmann T, Anaturk M, Suri S, Ebmeier KP et al. The maternal brain: Region-specific patterns of brain aging are traceable decades after childbirth. Hum Brain Mapp 2020; 41(16): 4718–4729.

19. Ning K, Zhao L, Franklin M, Matloff W, Batta I, Arzouni N et al. Parity is associated with cognitive function and brain age in both females and males. Sci Rep 2020; 10(1): 6100.

20. Orchard ER, Ward PGD, Sforazzini F, Storey E, Egan GF, Jamadar SD. Relationship between parenthood and cortical thickness in late adulthood. PLoS One 2020; 15(7): e0236031.

21. Read SL, Grundy EMD. Fertility History and Cognition in Later Life. J Gerontol B Psychol Sci Soc Sci 2017; 72(6): 1021–1031.

22. Tearne JE. Older maternal age and child behavioral and cognitive outcomes: a review of the literature. Fertil Steril 2015; 103(6): 1381–1391.

23. Kang JH, Grodstein F. Postmenopausal hormone therapy, timing of initiation, APOE and cognitive decline. Neurobiol Aging 2012; 33(7): 1129–1137.

24. Schmidt R, Fazekas F, Reinhart B, Kapeller P, Fazekas G, Offenbacher H et al. Estrogen replacement therapy in older women: a neuropsychological and brain MRI study. J Am Geriatr Soc 1996; 44(11): 1307–1313.

25. Kampen DL, Sherwin BB. Estrogen use and verbal memory in healthy postmenopausal women. Obstet Gynecol 1994; 83(6): 979–983.

26. Zec RF, Trivedi MA. The effects of estrogen replacement therapy on neuropsychological functioning in postmenopausal women with and without dementia: a critical and theoretical review. Neuropsychol Rev 2002; 12(2): 65–109.

27. Eisenberg DT, Kuzawa CW, Hayes MG. Worldwide allele frequencies of the human apolipoprotein E gene: climate, local adaptations, and evolutionary history. Am J Phys Anthropol 2010; 143(1): 100–111.

28. Yaffe K, Haan M, Byers A, Tangen C, Kuller L. Estrogen use, APOE, and cognitive decline: evidence of gene-environment interaction. Neurology 2000; 54(10): 1949–1954.

29. Dunkin J, Rasgon N, Wagner-Steh K, David S, Altshuler L, Rapkin A. Reproductive events modify the effects of estrogen replacement therapy on cognition in healthy postmenopausal women. Psychoneuroendocrino 2005; 30(3): 284–296.

30. Kang JH, Weuve J, Grodstein F. Postmenopausal hormone therapy and risk of cognitive decline in community-dwelling aging women. Neurology 2004; 63(1): 101–107.

31. MacLennan AH, Henderson VW, Paine BJ, Mathias J, Ramsay EN, Ryan P et al. Hormone therapy, timing of initiation, and cognition in women aged older than 60 years: the REMEMBER pilot study. Menopause 2006; 13(1): 28–36.

32. Maki PM, Sundermann E. Hormone therapy and cognitive function. Hum Reprod Update 2009; 15(6): 667–681.

33. Shao H, Breitner JC, Whitmer RA, Wang J, Hayden K, Wengreen H et al. Hormone therapy and Alzheimer disease dementia: new findings from the Cache County Study. Neurology 2012; 79(18): 1846–1852.

34. Gogos A. Natural and synthetic sex hormones: effects on higher-order cognitive function and prepulse inhibition. Biol Psychol 2013; 93(1): 17–23.

35. Mordecai KL, Rubin LH, Maki PM. Effects of menstrual cycle phase and oral contraceptive use on verbal memory. Horm Behav 2008; 54(2): 286–293.

36. Warren AM, Gurvich C, Worsley R, Kulkarni J. A systematic review of the impact of oral contraceptives on cognition. Contraception 2014; 90(2): 111–116.

37. Egan KR, Gleason CE. Longer duration of hormonal contraceptive use predicts better cognitive outcomes later in life. J Womens Health (Larchmt) 2012; 21(12): 1259–1266.

38. Lyall DM, Cullen B, Allerhand M, Smith DJ, Mackay D, Evans J et al. Cognitive Test Scores in UK Biobank: Data Reduction in 480,416 Participants and Longitudinal Stability in 20,346 Participants. PLoS One 2016; 11(4): e0154222.

39. Fawns-Ritchie C, Deary IJ. Reliability and validity of the UK Biobank cognitive tests. PLoS One 2020; 15(4): e0231627.

40. Bycroft C, Freeman C, Petkova D, Band G, Elliott LT, Sharp K et al. The UK Biobank resource with deep phenotyping and genomic data. Nature 2018; 562(7726): 203–209.

41. Lyall DM, Ward J, Ritchie SJ, Davies G, Cullen B, Celis C et al. Alzheimer disease genetic risk factor APOE e4 and cognitive abilities in 111,739 UK Biobank participants. Age Ageing 2016; 45(4): 511–517.

42. Lyall DM, Cox SR, Lyall LM, Celis-Morales C, Cullen B, Mackay DF et al. Association between APOE e4 and white matter hyperintensity volume, but not total brain volume or white matter integrity. Brain Imaging Behav 2020; 14(5): 1468–1476.

43. Wisdom NM, Callahan JL, Hawkins KA. The effects of apolipoprotein E on non-impaired cognitive functioning: a meta-analysis. Neurobiol Aging 2011; 32(1): 63–74.

44. Lovden M, Fratiglioni L, Glymour MM, Lindenberger U, Tucker-Drob EM. Education and Cognitive Functioning Across the Life Span. Psychol Sci Public Interest 2020; 21(1): 6–41.

45. Park DC, Reuter-Lorenz P. The adaptive brain: aging and neurocognitive scaffolding. Annu Rev Psychol 2009; 60: 173–196.

46. Boyle CP, Raji CA, Erickson KI, Lopez OL, Becker JT, Gach HM et al. Estrogen, brain structure, and cognition in postmenopausal women. Hum Brain Mapp 2021; 42(1): 24–35.

47. Team RC. R: A language and environment for statistical computing. 2013.

48. Nakagawa S, Cuthill IC. Effect size, confidence interval and statistical significance: a practical guide for biologists. Biol Rev Camb Philos Soc 2007; 82(4): 591–605.

49. Koo MM, Rohan TE. Accuracy of short-term recall of age at menarche. Ann Hum Biol 1997; 24(1): 61–64.

50. Kuh D, Butterworth S, Kok H, Richards M, Hardy R, Wadsworth ME et al. Childhood cognitive ability and age at menopause: evidence from two cohort studies. Menopause 2005; 12(4): 475–482.

51. Georgakis MK, Beskou-Kontou T, Theodoridis I, Skalkidou A, Petridou ET. Surgical menopause in association with cognitive function and risk of dementia: A systematic review and meta-analysis. Psychoneuroendocrino 2019; 106: 9–19.

52. Gong J, Harris K, Peters SAE, Woodward M. Reproductive factors and the risk of incident dementia: A cohort study of UK Biobank participants. PLoS Med 2022; 19(4): e1003955.

53. Ley SH, Li Y, Tobias DK, Manson JE, Rosner B, Hu FB et al. Duration of Reproductive Life Span, Age at Menarche, and Age at Menopause Are Associated With Risk of Cardiovascular Disease in Women. J Am Heart Assoc 2017; 6(11).

54. Thomas MP, Potter BV. The structural biology of oestrogen metabolism. J Steroid Biochem Mol Biol 2013; 137: 27–49.

55. al-Azzawi F. Endocrinological aspects of the menopause. Br Med Bull 1992; 48(2): 262–275.

56. Cardenas EF, Kujawa A, Humphreys KL. Neurobiological changes during the peripartum period: implications for health and behavior. Soc Cogn Affect Neurosci 2020; 15(10): 1097–1110.

57. Barnes A. Low fertility: a discussion paper. FaHCSIA Occasional Paper 2001; (2).

58. Henry JD, Rendell PG. A review of the impact of pregnancy on memory function. J Clin Exp Neuropsychol 2007; 29(8): 793–803.

59. Rentz DM, Weiss BK, Jacobs EG, Cherkerzian S, Klibanski A, Remington A et al. Sex differences in episodic memory in early midlife: impact of reproductive aging. Menopause 2017; 24(4): 400–408.

60. Natri H, Garcia AR, Buetow KH, Trumble BC, Wilson MA. The Pregnancy Pickle: Evolved Immune Compensation Due to Pregnancy Underlies Sex Differences in Human Diseases. Trends Genet 2019; 35(7): 478–488.

61. Fox M, Berzuini C, Knapp LA, Glynn LM. Women’s Pregnancy Life History and Alzheimer’s Risk: Can Immunoregulation Explain the Link? Am J Alzheimers Dis Other Demen 2018; 33(8): 516–526.

62. Barba-Muller E, Craddock S, Carmona S, Hoekzema E. Brain plasticity in pregnancy and the postpartum period: links to maternal caregiving and mental health. Arch Womens Ment Health 2019; 22(2): 289–299.

63. Diamond A. Executive functions. Annu Rev Psychol 2013; 64: 135–168.

64. Kim P. Human Maternal Brain Plasticity: Adaptation to Parenting. New Dir Child Adolesc Dev 2016; 2016(153): 47–58.

65. Berent-Spillson A, Briceno E, Pinsky A, Simmen A, Persad CC, Zubieta JK et al. Distinct cognitive effects of estrogen and progesterone in menopausal women. Psychoneuroendocrino 2015; 59: 25–36.

66. Maki PM. Critical window hypothesis of hormone therapy and cognition: a scientific update on clinical studies. Menopause 2013; 20(6): 695–709.

67. Matthews KA, Kuller LH, Wing RR, Meilahn EN, Plantinga P. Prior to use of estrogen replacement therapy, are users healthier than nonusers? American journal of epidemiology 1996; 143(10): 971–978.

68. Barrett-Connor E, Laughlin GA. Endogenous and exogenous estrogen, cognitive function, and dementia in postmenopausal women: evidence from epidemiologic studies and clinical trials. Semin Reprod Med 2009; 27(3): 275–282.

69. Hampson E. A brief guide to the menstrual cycle and oral contraceptive use for researchers in behavioral endocrinology. Hormones and Behavior 2020; 119: 104655.

