## Supplementary Materials for "Female-specific factors are associated with cognition in the UK Biobank cohort"

***Note 1: Participants with diagnosis known to influence cognition***

Participants with diagnosed brain disorders known to influence cognition were excluded from the main sample (n = 39,011). Brain disorders were based on International Classification of Diseases (ICD)-10 diagnoses (chapter V and VI, field F; mental and behavioral disorders), including F00-F03 for Alzheimer’s disease and dementia, and F06.7 (´Mild cognitive disorder`), field G (`Diseases of the nervous system´), including inflammatory and neurodegenerative diseases (except G55-59; ´Nerve, nerve root and plexus disorders´) and field I (‘Diseases of the circulatory system’), including I64 for stroke. An overview of the diagnoses is provided in the UK Biobank online resources (<https://biobank.ndph.ox.ac.uk/showcase/field.cgi?id=41270>), and the diagnostic criteria are listed in the ICD10 diagnostic manual (<https://www.who.int/classifications/icd/icdonlineversions>).

***Note 2: Townsend deprivation index and lifestyle score***

The Townsend deprivation index was derived from national census data about car ownership, household overcrowding, owner occupation, and unemployment aggregated for postcodes of residence in the UK^1^. Higher scores reflect higher levels of socioeconomic deprivation. The lifestyle score was calculated based on sleep duration, time spent watching television, current and past smoking status, alcohol consumption frequency, physical activity level, intake of fruits and vegetables, and intake of oily fish, beef, lamb/mutton, pork and processed meat^1^. Each unhealthy lifestyle factor was scored with one point (e.g., smoking), and each participant’s points were summed to generate an unweighted score (from 0-9): the higher the lifestyle score, the unhealthier the participant’s lifestyle.

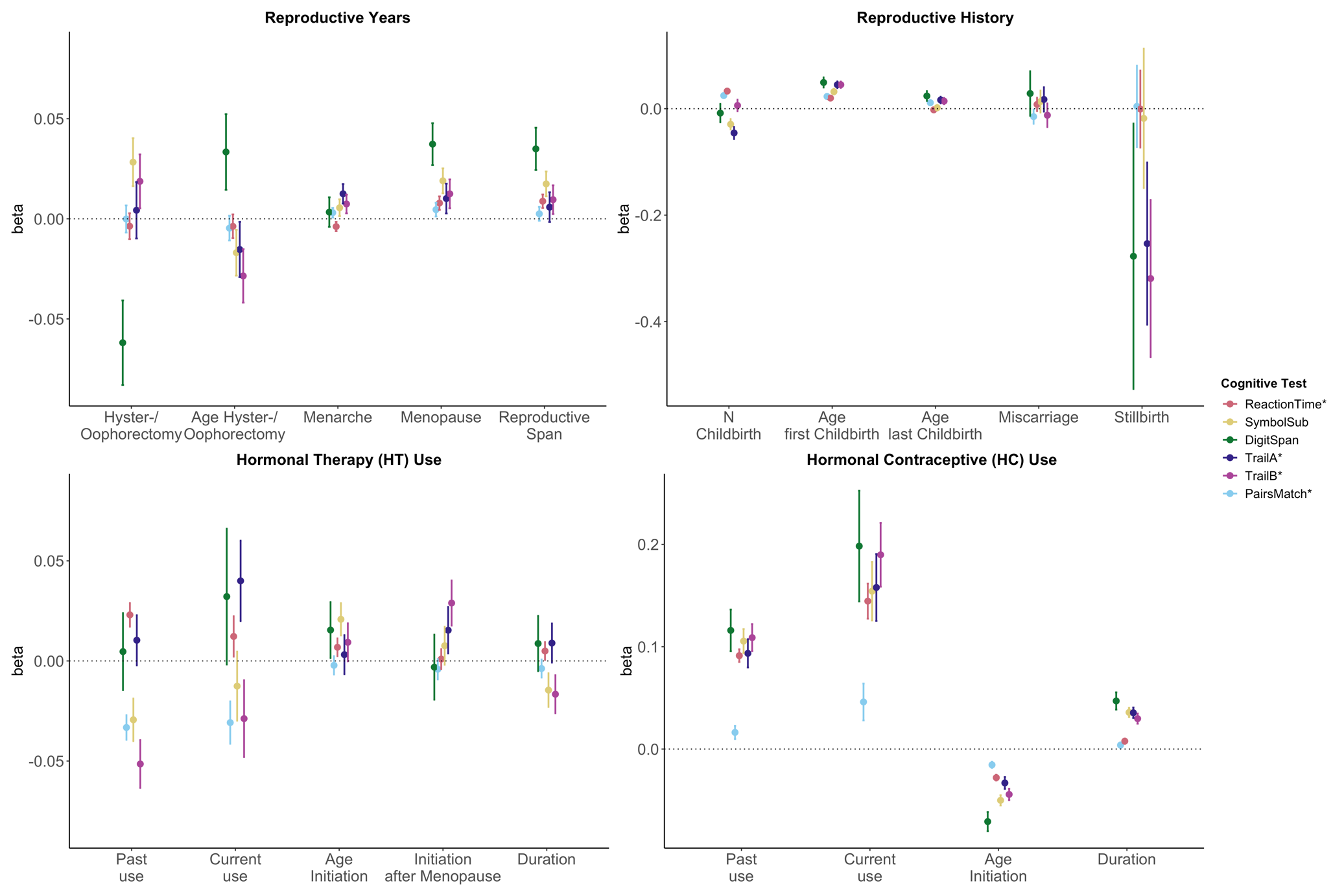

**Figure S1| Associations between female-specific factors and cognitive performance.**

Point plot of beta-values with standard error from separate multiple regression analysis with cognitive task as dependent variable and female-specific variables as independent variable. All models are adjusted for age, education, body mass index, Townsend deprivation score, lifestyle score. In addition, the analyses for reproductive span and age at menopause were corrected for use of HT, use of HC, history of hysterectomy and bilateral oophorectomy, and number of live births. The HT models were additionally adjusted for history of hysterectomy and bilateral oophorectomy, and the hysterectomy/oophorectomy model was co-varied for use of HT. All variables were standardized prior to performing the multiple linear regression analysis (subtracting the mean and dividing by the standard deviation). For visualization purposes, cognitive tests marked with * are inverted (multiplied by -1) so that positive beta-values always indicate associations between higher values on the female-specific variables and better performance on cognitive tests.

***Table S1| Sample demographics stratified by hormone therapy (HT) user groups.***

|  | **Never-User** | **Current-User** | **Past-User** | **p-value** | **test** |
| --- | --- | --- | --- | --- | --- |
| **Total N** | 138,963 | 12,663 | 60,083 |  |  |
| **Age** (years)* | 53.5 ± 8.0 | 56.7 ± 6.3 | 61.2 ± 5.1 | <0.001 | KW |
| **Age range** (years) | 39 – 70 | 40 – 70 | 40 – 70 |  |  |
| **Education**, N (%) |  |  |  | <0.001 | χ2 |
| College/University degree | 51,475 (37.3) | 4,343 (34.7) | 16,020 (26.9) |  |  |
| O levels/GCSEs or equivalent | 32,214 (23.3) | 3,017 (24.1) | 14,620 (24.5) |  |  |
| None of the above | 15,542 (11.3) | 1,580 (12.6) | 12,965 (21.7) |  |  |
| A levels/AS levels or equivalent | 18,609 (13.5) | 1,561 (12.5) | 6,233 (10.5) |  |  |
| Other professional qualifications | 6,766 (4.9) | 814 (6.5) | 4,562 (7.7) |  |  |
| CSEs or equivalent | 8,120 (5.9) | 673 (5.4) | 2,320 (3.9) |  |  |
| NVQ/HND/HNC or equivalent | 5,337 (3.9) | 537 (4.3) | 2,912 (4.9) |  |  |
| **Townsend Deprivation Score*** | -1.5 ± 3.0 | -1.6 ± 2.9 | -1.7 ± 2.9 | <0.001 | KW |
| **Lifestyle score*** | 2.9 ± 1.6 | 3.0 ± 1.6 | 2.9 ± 1.6 | <0.001 | KW |
| **BMI** (kg/m^2^) | 26.7 ± 5.1 | 26.3 ± 4.5 | 27.1 ± 4.7 | <0.001 | KW |
| **Bilateral Oophorectomy**, Yes N (%) | 3,151 (2.3) | 3,135 (24.8) | 9,458 (15.7) | <0.001 | χ2 |
| **Hysterectomy**, Yes N (%) | 4,241 (3.2) | 1,325 (14.9) | 7,540 (15.2) | <0.001 | χ2 |
| **Age first used HT***^x^ (years) |  | 47.1 ± 5.9 | 47.8 ± 5.1 | <0.001 | KW |
| **Age last used HT***^x^ (years) |  | 56.6 ± 6.3 | 54.1 ± 5.7 | <0.001 | KW |
| **Duration HT use***^x^ (years) |  | 9.4 ± 6.8 | 6.3 ± 5.2 | <0.001 | KW |

* Continuous data in mean ± standard deviation and categorical data as number (%).

***^x^*** Complete data for 12,270 current-users and 58,467 past-users.

Abbreviations: N = Number; O = Ordinary Level Qualification; GCSE = General Certificate of Secondary Education; A = Advanced Level Qualification; AS = Advanced Subsidiary Level Qualification; CSE = Certificate of Secondary Education; NVQ = National Vocational Qualification; HND = Higher National Diploma; HNC = Higher National Certificate; KW = Kruskal-Wallis.

***Table S2| Sample demographics stratified by hormonal contraceptive (HC) user groups.***

|  | **Never-User** | **Current-User** | **Past-User** | **p-value** | **test** |
| --- | --- | --- | --- | --- | --- |
| **Total N** | 39,908 | 4,322 | 160,392 |  |  |
| **Age** (years)* | 60.0 ± 7.8 | 45.6 ± 4.0 | 55.3 ± 7.7 | <0.001 | KW |
| **Age range** (years) | 39 – 70 | 40 – 70 | 40 – 70 |  |  |
| **Education**, N (%) |  |  |  | <0.001 | χ2 |
| College/University degree | 11,215 (28.3) | 1,742 (40.7) | 56,262 (35.3) |  |  |
| O levels/GCSEs or equivalent | 8,695 (22.0) | 1,100 (25.7) | 38,212 (24.0) |  |  |
| None of the above | 9,794 (24.7) | 116 (2.7) | 19,761 (12.4) |  |  |
| A levels/AS levels or equivalent | 3,863 (9.8) | 747 (17.4) | 20,675 (13.0) |  |  |
| Other professional qualifications | 2,851 (7.2) | 109 (2.5) | 8,882 (5.6) |  |  |
| CSEs or equivalent | 1,440 (3.6) | 333 (7.8) | 8,844 (5.6) |  |  |
| NVQ/HND/HNC or equivalent | 1,737 (4.4) | 136 (3.2) | 6,680 (4.2) |  |  |
| **Townsend Deprivation Score*** | -1.3 ± 3.0 | -1.3 ± 3.0 | -1.6 ± 2.9 | <0.001 | KW |
| **Lifestyle score*** | 2.8 ± 1.6 | 3.1 ± 1.6 | 2.9 ± 1.6 | <0.001 | KW |
| **BMI** (kg/m^2^) | 27.2 ± 5.2 | 25.8 ± 4.7 | 26.8 ± 4.9 | <0.001 | KW |
| **Age first used HC***^x^ |  | 20.3 ± 5.6 | 21.5 ± 4.6 | <0.001 | KW |
| **Age last used HC***^x^ (years) |  | 45.6 ± 4.0 | 31.9 ± 7.5 | <0.001 | KW |
| **Duration HC use***^x^ |  | 25.2 ± 6.6 | 10.4 ± 7.7 | <0.001 | KW |

* Continuous data in mean ± standard deviation and categorical data as number (%).

***^x^*** Complete data for 4,219 current-users and 158,920 past-users.

Abbreviations: N = Number; O = Ordinary Level Qualification; GCSE = General Certificate of Secondary Education; A = Advanced Level Qualification; AS = Advanced Subsidiary Level Qualification; CSE = Certificate of Secondary Education; NVQ = National Vocational Qualification; HND = Higher National Diploma; HNC = Higher National Certificate; KW = Kruskal-Wallis.

***Table S3| Sample demographics stratified by APOE*** ε***4 status.***

|  | **Non-Carrier** | **Carrier** | **p-value** | **test** |
| --- | --- | --- | --- | --- |
| **Total N** | **153,395** | **55,423** |  |  |
| **Age** (years)* | 57.0 [50.0, 63.0] | 57.0 [50.0, 63.0] | <0.001 | KW |
| **Education**, N (%) |  |  | 0.039 | χ2 |
| College/University degree | 51,463 (33.6) | 18,497 (33.4) |  |  |
| O levels/GCSEs or equivalent | 36,225 (23.6) | 13,369 (24.1) |  |  |
| None of the above | 23,253 (15.2) | 8,168 (14.7) |  |  |
| A levels/AS levels or equivalent | 18,949 (12.4) | 6,921 (12.5) |  |  |
| Other professional qualifications | 8,951 (5.8) | 3,143 (5.7) |  |  |
| CSEs or equivalent | 7,991 (5.2) | 2,897 (5.2) |  |  |
| NVQ/HND/HNC or equivalent | 6,423 (4.2) | 2,382 (4.3) |  |  |
| **Townsend Deprivation Score*** | -2.3 [-3.7, 0.1] | -2.3 [-3.7, 0.1] | 0.003 | KW |
| **Lifestyle score*** | 3.0 [2.0, 4.0] | 3.0 [2.0, 4.0] | <0.001 | KW |
| **BMI** (kg/m^2^) | 25.9 [23.3, 29.4] | 25.8 [23.3, 29.3] | <0.001 | KW |
| **APOE e4 alleles**, N (%) |  |  |  |  |
| 1 x e4 allele |  | 50458 (91.0) |  |  |
| 2 x e4 allele |  | 4965 (9.0) |  |  |

*Continuous data in median [Interquartile range] and categorical data as number (%).

Abbreviations: N = Number; O = Ordinary Level Qualification; GCSE = General Certificate of Secondary Education; A = Advanced Level Qualification; AS = Advanced Subsidiary Level Qualification; CSE = Certificate of Secondary Education; NVQ = National Vocational Qualification; HND = Higher National Diploma; HNC = Higher National Certificate; KW = Kruskal-Wallis.

***Table S4| Associations between female-specific factors and late life cognition.***

| **Model** | **Test** | **beta** | **S.E.** | **t** | **p** | **pFDR** | **d** |
| --- | --- | --- | --- | --- | --- | --- | --- |
| Reproductive Span | PairsMatch | 0.003 | 0.003 | 0.731 | 0.465 | 0.586 | 0.005 |
|  | ReactionTime | 0.009 | 0.003 | 2.619 | **0.009** | **0.025** | 0.017 |
|  | SymbolSub | 0.017 | 0.006 | 2.817 | **0.005** | **0.016** | 0.035 |
|  | DigitSpan | 0.035 | 0.011 | 3.309 | **0.001** | **0.003** | 0.066 |
|  | TrailA | 0.006 | 0.007 | 0.784 | 0.433 | 0.566 | 0.010 |
|  | TrailB | 0.010 | 0.007 | 1.333 | 0.182 | 0.319 | 0.018 |
| Age at Menarche | PairsMatch | 0.003 | 0.002 | 1.271 | 0.204 | 0.336 | 0.006 |
|  | ReactionTime | -0.004 | 0.002 | -1.675 | 0.094 | 0.191 | -0.008 |
|  | SymbolSub | 0.006 | 0.004 | 1.322 | 0.186 | 0.319 | 0.012 |
|  | DigitSpan | 0.003 | 0.007 | 0.456 | 0.649 | 0.702 | 0.007 |
|  | TrailA | 0.013 | 0.005 | 2.560 | **0.010** | **0.028** | 0.025 |
|  | TrailB | 0.007 | 0.005 | 1.591 | 0.112 | 0.220 | 0.016 |
| Age at Menopause | PairsMatch | 0.005 | 0.003 | 1.339 | 0.181 | 0.319 | 0.009 |
|  | ReactionTime | 0.008 | 0.003 | 2.346 | **0.019** | **0.046** | 0.015 |
|  | SymbolSub | 0.019 | 0.006 | 3.062 | **0.002** | **0.008** | 0.037 |
|  | DigitSpan | 0.037 | 0.010 | 3.564 | **3.67e-04** | **0.001** | 0.071 |
|  | TrailA | 0.010 | 0.007 | 1.365 | 0.172 | 0.319 | 0.018 |
|  | TrailB | 0.012 | 0.007 | 1.737 | 0.082 | 0.172 | 0.023 |
| Oophorectomy | PairsMatch | 0.009 | 0.009 | 0.941 | 0.347 | 0.487 | 0.009 |
|  | ReactionTime | -0.017 | 0.009 | -1.846 | 0.065 | 0.141 | -0.017 |
|  | SymbolSub | 0.036 | 0.017 | 2.120 | **0.034** | 0.079 | 0.040 |
|  | DigitSpan | -0.038 | 0.029 | -1.308 | 0.191 | 0.319 | -0.037 |
|  | TrailA | 0.023 | 0.020 | 1.178 | 0.239 | 0.371 | 0.024 |
|  | TrailB | 0.012 | 0.019 | 0.622 | 0.534 | 0.618 | 0.013 |
| Age at Oophorectomy | PairsMatch | -0.013 | 0.010 | -1.241 | 0.215 | 0.346 | -0.022 |
|  | ReactionTime | -0.006 | 0.010 | -0.597 | 0.550 | 0.632 | -0.011 |
|  | SymbolSub | -0.003 | 0.019 | -0.164 | 0.870 | 0.897 | -0.006 |
|  | DigitSpan | 0.032 | 0.029 | 1.104 | 0.270 | 0.397 | 0.061 |
|  | TrailA | -0.028 | 0.023 | -1.228 | 0.220 | 0.349 | -0.049 |
|  | TrailB | -0.049 | 0.023 | -2.161 | **0.031** | 0.073 | -0.086 |
| Hysterectomy | PairsMatch | -0.006 | 0.009 | -0.716 | 0.474 | 0.590 | -0.006 |
|  | ReactionTime | 0.005 | 0.008 | 0.650 | 0.515 | 0.613 | 0.006 |
|  | SymbolSub | 0.020 | 0.015 | 1.329 | 0.184 | 0.319 | 0.023 |
|  | DigitSpan | -0.071 | 0.027 | -2.648 | **0.008** | **0.025** | -0.072 |
|  | TrailA | -0.009 | 0.018 | -0.502 | 0.616 | 0.683 | -0.009 |
|  | TrailB | 0.021 | 0.017 | 1.254 | 0.210 | 0.342 | 0.023 |
| Age at Hysterectomy | PairsMatch | -0.011 | 0.008 | -1.354 | 0.176 | 0.319 | -0.022 |
|  | ReactionTime | 0.004 | 0.008 | 0.513 | 0.608 | 0.680 | 0.008 |
|  | SymbolSub | -0.033 | 0.016 | -2.085 | **0.037** | 0.084 | -0.068 |
|  | DigitSpan | 0.002 | 0.026 | 0.087 | 0.931 | 0.945 | 0.005 |
|  | TrailA | -0.025 | 0.019 | -1.311 | 0.190 | 0.319 | -0.046 |
|  | TrailB | -0.018 | 0.018 | -0.993 | 0.321 | 0.465 | -0.035 |
| N Childbirth | PairsMatch | 0.025 | 0.004 | 5.628 | **1.82e-08** | **1.05e-07** | 0.027 |
|  | ReactionTime | 0.033 | 0.004 | 7.785 | **6.99e-15** | **1.03e-13** | 0.037 |
|  | SymbolSub | -0.029 | 0.010 | -2.978 | **0.003** | **0.010** | -0.027 |
|  | DigitSpan | -0.008 | 0.017 | -0.474 | 0.636 | 0.693 | -0.007 |
|  | TrailA | -0.046 | 0.011 | -4.002 | **6.29e-05** | **2.68e-04** | -0.039 |
|  | TrailB | 0.006 | 0.011 | 0.555 | 0.579 | 0.655 | 0.005 |
| N Childbirth^2^ | PairsMatch | -0.011 | 0.004 | -2.533 | **0.011** | **0.029** | -0.012 |
|  | ReactionTime | -0.033 | 0.004 | -7.631 | **2.34e-14** | **3.09e-13** | -0.036 |
|  | SymbolSub | -0.011 | 0.012 | -0.888 | 0.374 | 0.515 | -0.008 |
|  | DigitSpan | -0.013 | 0.019 | -0.694 | 0.488 | 0.601 | -0.010 |
|  | TrailA | -0.010 | 0.014 | -0.665 | 0.506 | 0.613 | -0.006 |
|  | TrailB | -0.028 | 0.014 | -2.059 | **0.039** | 0.088 | -0.020 |
| Age last Birth | PairsMatch | 0.011 | 0.003 | 3.780 | **1.57e-04** | **0.001** | 0.022 |
|  | ReactionTime | -0.002 | 0.003 | -0.741 | 0.459 | 0.586 | -0.004 |
|  | SymbolSub | 0.002 | 0.005 | 0.350 | 0.726 | 0.779 | 0.004 |
|  | DigitSpan | 0.024 | 0.009 | 2.618 | **0.009** | **0.025** | 0.047 |
|  | TrailA | 0.016 | 0.006 | 2.642 | **0.008** | **0.025** | 0.032 |
|  | TrailB | 0.014 | 0.006 | 2.431 | **0.015** | **0.037** | 0.030 |
| Age first Birth | PairsMatch | 0.023 | 0.003 | 7.443 | **9.90e-14** | **1.01e-12** | 0.043 |
|  | ReactionTime | 0.020 | 0.003 | 6.727 | **1.74e-11** | **1.35e-10** | 0.039 |
|  | SymbolSub | 0.032 | 0.005 | 6.001 | **1.98e-09** | **1.31e-08** | 0.068 |
|  | DigitSpan | 0.049 | 0.009 | 5.209 | **1.93e-07** | **9.44e-07** | 0.094 |
|  | TrailA | 0.045 | 0.006 | 7.202 | **6.11e-13** | **5.76e-12** | 0.088 |
|  | TrailB | 0.045 | 0.006 | 7.593 | **3.24e-14** | **3.56e-13** | 0.093 |
| Miscarriage & Termination | PairsMatch | -0.015 | 0.013 | -1.102 | 0.271 | 0.397 | -0.015 |
|  | ReactionTime | 0.008 | 0.013 | 0.631 | 0.528 | 0.617 | 0.009 |
|  | SymbolSub | 0.013 | 0.021 | 0.654 | 0.513 | 0.613 | 0.015 |
|  | DigitSpan | 0.029 | 0.042 | 0.684 | 0.494 | 0.603 | 0.030 |
|  | TrailA | 0.017 | 0.023 | 0.761 | 0.447 | 0.578 | 0.019 |
|  | TrailB | -0.012 | 0.022 | -0.553 | 0.580 | 0.655 | -0.014 |
| Stillbirth | PairsMatch | 0.004 | 0.077 | 0.058 | 0.954 | 0.961 | 0.005 |
|  | ReactionTime | -0.001 | 0.072 | -0.009 | 0.993 | 0.993 | -0.001 |
|  | SymbolSub | -0.018 | 0.131 | -0.138 | 0.890 | 0.911 | -0.021 |
|  | DigitSpan | -0.277 | 0.249 | -1.111 | 0.266 | 0.397 | -0.288 |
|  | TrailA | -0.254 | 0.152 | -1.663 | 0.096 | 0.193 | -0.274 |
|  | TrailB | -0.319 | 0.148 | -2.161 | **0.031** | 0.073 | -0.356 |
| Current HT use | PairsMatch | -0.031 | 0.011 | -2.911 | **0.004** | **0.012** | -0.026 |
|  | ReactionTime | 0.012 | 0.010 | 1.212 | 0.225 | 0.354 | 0.011 |
|  | SymbolSub | -0.013 | 0.017 | -0.729 | 0.466 | 0.586 | -0.012 |
|  | DigitSpan | 0.032 | 0.034 | 0.947 | 0.344 | 0.487 | 0.016 |
|  | TrailA | 0.040 | 0.020 | 1.993 | **0.046** | 0.102 | 0.022 |
|  | TrailB | -0.029 | 0.019 | -1.501 | 0.133 | 0.255 | -0.017 |
| Past HT use | PairsMatch | -0.033 | 0.006 | -5.394 | **6.89e-08** | **3.64e-07** | -0.048 |
|  | ReactionTime | 0.023 | 0.006 | 3.911 | **9.19e-05** | **3.79e-04** | 0.036 |
|  | SymbolSub | -0.029 | 0.011 | -2.763 | **0.006** | **0.018** | -0.046 |
|  | DigitSpan | 0.005 | 0.019 | 0.242 | 0.809 | 0.854 | 0.004 |
|  | TrailA | 0.010 | 0.013 | 0.826 | 0.409 | 0.550 | 0.009 |
|  | TrailB | -0.051 | 0.012 | -4.299 | **1.72e-05** | **7.56e-05** | -0.048 |
| Age HT initiation | PairsMatch | -0.002 | 0.005 | -0.478 | 0.633 | 0.693 | -0.004 |
|  | ReactionTime | 0.007 | 0.004 | 1.546 | 0.122 | 0.237 | 0.013 |
|  | SymbolSub | 0.021 | 0.008 | 2.588 | **0.010** | **0.027** | 0.042 |
|  | DigitSpan | 0.015 | 0.014 | 1.105 | 0.269 | 0.397 | 0.029 |
|  | TrailA | 0.003 | 0.010 | 0.323 | 0.747 | 0.795 | 0.006 |
|  | TrailB | 0.009 | 0.009 | 0.982 | 0.326 | 0.468 | 0.017 |
| Age HT rel. Menopause | PairsMatch | -0.004 | 0.005 | -0.815 | 0.415 | 0.554 | -0.008 |
|  | ReactionTime | 0.001 | 0.005 | 0.188 | 0.851 | 0.885 | 0.002 |
|  | SymbolSub | 0.008 | 0.009 | 0.802 | 0.422 | 0.558 | 0.016 |
|  | DigitSpan | -0.003 | 0.016 | -0.191 | 0.848 | 0.885 | -0.006 |
|  | TrailA | 0.015 | 0.012 | 1.323 | 0.186 | 0.319 | 0.028 |
|  | TrailB | 0.029 | 0.011 | 2.562 | **0.010** | **0.028** | 0.054 |
| Duration HT use | PairsMatch | -0.004 | 0.005 | -0.836 | 0.403 | 0.548 | -0.007 |
|  | ReactionTime | 0.005 | 0.004 | 1.112 | 0.266 | 0.397 | 0.009 |
|  | SymbolSub | -0.015 | 0.008 | -1.732 | 0.083 | 0.172 | -0.028 |
|  | DigitSpan | 0.009 | 0.014 | 0.631 | 0.528 | 0.617 | 0.017 |
|  | TrailA | 0.009 | 0.010 | 0.915 | 0.360 | 0.501 | 0.016 |
|  | TrailB | -0.017 | 0.009 | -1.751 | 0.080 | 0.170 | -0.031 |
| Current HC use | PairsMatch | 0.046 | 0.018 | 2.558 | **0.011** | **0.028** | 0.021 |
|  | ReactionTime | 0.145 | 0.017 | 8.440 | **3.19e-17** | **8.41e-16** | 0.059 |
|  | SymbolSub | 0.154 | 0.029 | 5.387 | **7.19e-08** | **3.65e-07** | 0.069 |
|  | DigitSpan | 0.198 | 0.054 | 3.666 | **2.47e-04** | **0.001** | 0.072 |
|  | TrailA | 0.158 | 0.033 | 4.836 | **1.33e-06** | **6.07e-06** | 0.068 |
|  | TrailB | 0.190 | 0.031 | 6.085 | **1.18e-09** | **8.64e-09** | 0.087 |
| Past HC use | PairsMatch | 0.016 | 0.006 | 2.521 | **0.012** | **0.030** | 0.020 |
|  | ReactionTime | 0.091 | 0.006 | 14.795 | **1.70e-49** | **2.25e-47** | 0.113 |
|  | SymbolSub | 0.105 | 0.012 | 9.023 | **1.89e-19** | **6.25e-18** | 0.116 |
|  | DigitSpan | 0.116 | 0.020 | 5.688 | **1.30e-08** | **7.83e-08** | 0.113 |
|  | TrailA | 0.094 | 0.014 | 6.763 | **1.37e-11** | **1.13e-10** | 0.089 |
|  | TrailB | 0.109 | 0.013 | 8.239 | **1.79e-16** | **3.93e-15** | 0.117 |
| Age HC initiation | PairsMatch | -0.015 | 0.003 | -5.195 | **2.05e-07** | **9.65e-07** | -0.028 |
|  | ReactionTime | -0.028 | 0.003 | -9.927 | **3.22e-23** | **1.42e-21** | -0.053 |
|  | SymbolSub | -0.050 | 0.005 | -10.117 | **4.94e-24** | **3.26e-22** | -0.100 |
|  | DigitSpan | -0.071 | 0.009 | -7.603 | **3.08e-14** | **3.56e-13** | -0.126 |
|  | TrailA | -0.033 | 0.006 | -5.699 | **1.21e-08** | **7.63e-08** | -0.060 |
|  | TrailB | -0.044 | 0.006 | -8.014 | **1.15e-15** | **1.89e-14** | -0.085 |
| Duration HC use | PairsMatch | 0.004 | 0.003 | 1.385 | 0.166 | 0.313 | 0.008 |
|  | ReactionTime | 0.008 | 0.003 | 2.934 | **0.003** | **0.011** | 0.016 |
|  | SymbolSub | 0.036 | 0.004 | 8.057 | **8.06e-16** | **1.52e-14** | 0.082 |
|  | DigitSpan | 0.047 | 0.008 | 5.596 | **2.23e-08** | **1.23e-07** | 0.095 |
|  | TrailA | 0.035 | 0.005 | 6.908 | **4.99e-12** | **4.39e-11** | 0.075 |
|  | TrailB | 0.030 | 0.005 | 6.045 | **1.51e-09** | **1.05e-08** | 0.066 |

Abbreviation: PairMatch = Pair Matching Test, ReactionTime = Reaction Time Test, SymbolSub = Symbol Substitution Test, DigitSpan = Digit Span Test, Trail A & B = Trail Making Test A & B, N = Number, HT = Hormone Therapy, HC = Hormonal Contraceptive, S.E. = Standard Error, FDR = False Discovery Rate. Significant results are highlighted in bold.

***Table S5| Non-linear effects of number of childbirths on cognitive functioning.***

|  | **Reaction Time** | | | | **Pair Matching** | | | |
| --- | --- | --- | --- | --- | --- | --- | --- | --- |
| **N Childbirth** | **beta** | **S.E.** | **t** | **p** | **beta** | **S.E.** | **t** | **p** |
| 0 (intercept) | -0.036 | 0.005 | -7.032 | **2.05e-12** | -0.032 | 0.005 | -5.848 | **4.99e-09** |
| 1 | 0.035 | 0.008 | 4.362 | **1.29e-05** | 0.021 | 0.008 | 2.498 | **0.0125** |
| 2 | 0.059 | 0.006 | 9.444 | **< 2e-16** | 0.031 | 0.007 | 4.767 | **1.87e-06** |
| 3 | 0.042 | 0.008 | 5.580 | **2.40e-08** | 0.051 | 0.008 | 6.524 | **6.87e-11** |
| 4 | -0.002 | 0.012 | -0.143 | 0.886 | 0.054 | 0.013 | 4.317 | **1.58e-05** |
| 5 | -0.066 | 0.024 | -2.719 | **0.007** | 0.025 | 0.026 | 0.963 | 0.336 |
| 6 | -0.107 | 0.046 | -2.341 | **0.019** | 0.022 | 0.047 | 0.467 | 0.640 |
| 7+ | -0.107 | 0.063 | 1.715 | 0.086 | 0.061 | 0.065 | 0.950 | 0.342 |

Abbreviation: N = Number, S.E. = Standard Error

***Table S6| Associations between female-specific factors and late life cognition after removal of extreme values.***

| **Model** | **Test** | **beta** | **S.E.** | **t** | **p** | **p_FDR_** | **d** |
| --- | --- | --- | --- | --- | --- | --- | --- |
| Reproductive Span | PairsMatch | 0.004 | 0.003 | 1.150 | 0.250 | 0.393 | 0.007 |
|  | ReactionTime | 0.008 | 0.003 | 2.455 | **0.014** | **0.038** | 0.016 |
|  | SymbolSub | 0.015 | 0.006 | 2.436 | **0.015** | **0.039** | 0.030 |
|  | DigitSpan | 0.037 | 0.010 | 3.538 | **4.05e-04** | **0.002** | 0.072 |
|  | TrailA | 0.008 | 0.007 | 1.077 | 0.281 | 0.413 | 0.014 |
|  | TrailB | 0.010 | 0.007 | 1.416 | 0.157 | 0.291 | 0.019 |
| Age at Menarche | PairsMatch | 0.003 | 0.002 | 1.364 | 0.173 | 0.312 | 0.007 |
|  | ReactionTime | -0.001 | 0.002 | -0.496 | 0.620 | 0.682 | -0.002 |
|  | SymbolSub | 0.007 | 0.004 | 1.598 | 0.110 | 0.220 | 0.015 |
|  | DigitSpan | 0.008 | 0.007 | 1.032 | 0.302 | 0.424 | 0.016 |
|  | TrailA | 0.013 | 0.005 | 2.614 | **0.009** | **0.028** | 0.026 |
|  | TrailB | 0.008 | 0.005 | 1.710 | 0.087 | 0.186 | 0.017 |
| Age at Menopause | PairsMatch | 0.005 | 0.003 | 1.570 | 0.117 | 0.230 | 0.010 |
|  | ReactionTime | 0.008 | 0.003 | 2.549 | **0.011** | **0.032** | 0.016 |
|  | SymbolSub | 0.017 | 0.006 | 2.763 | **0.006** | **0.019** | 0.034 |
|  | DigitSpan | 0.041 | 0.010 | 3.955 | **7.70e-05** | **3.18e-04** | 0.079 |
|  | TrailA | 0.012 | 0.007 | 1.722 | 0.085 | 0.184 | 0.023 |
|  | TrailB | 0.012 | 0.007 | 1.701 | 0.089 | 0.186 | 0.023 |
| Oophorectomy | PairsMatch | 0.009 | 0.009 | 0.941 | 0.347 | 0.477 | 0.009 |
|  | ReactionTime | -0.017 | 0.009 | -1.846 | 0.065 | 0.148 | -0.017 |
|  | SymbolSub | 0.036 | 0.017 | 2.120 | **0.034** | 0.085 | 0.040 |
|  | DigitSpan | -0.038 | 0.029 | -1.308 | 0.191 | 0.327 | -0.037 |
|  | TrailA | 0.023 | 0.020 | 1.178 | 0.239 | 0.379 | 0.024 |
|  | TrailB | 0.012 | 0.019 | 0.622 | 0.534 | 0.623 | 0.013 |
| Age at Oophorectomy | PairsMatch | -0.011 | 0.010 | -1.106 | 0.269 | 0.406 | -0.020 |
|  | ReactionTime | -0.008 | 0.010 | -0.765 | 0.444 | 0.574 | -0.014 |
|  | SymbolSub | 0.000 | 0.019 | 0.025 | 0.980 | 0.987 | 0.001 |
|  | DigitSpan | 0.031 | 0.029 | 1.061 | 0.289 | 0.414 | 0.059 |
|  | TrailA | -0.018 | 0.023 | -0.771 | 0.441 | 0.574 | -0.031 |
|  | TrailB | -0.040 | 0.023 | -1.772 | 0.076 | 0.168 | -0.070 |
| Hysterectomy | PairsMatch | -0.006 | 0.009 | -0.716 | 0.474 | 0.582 | -0.006 |
|  | ReactionTime | 0.005 | 0.008 | 0.650 | 0.515 | 0.613 | 0.006 |
|  | SymbolSub | 0.020 | 0.015 | 1.329 | 0.184 | 0.327 | 0.023 |
|  | DigitSpan | -0.071 | 0.027 | -2.648 | **0.008** | **0.026** | -0.072 |
|  | TrailA | -0.009 | 0.018 | -0.502 | 0.616 | 0.682 | -0.009 |
|  | TrailB | 0.021 | 0.017 | 1.254 | 0.210 | 0.355 | 0.023 |
| Age at Hysterectomy | PairsMatch | -0.010 | 0.008 | -1.234 | 0.217 | 0.363 | -0.020 |
|  | ReactionTime | 0.003 | 0.008 | 0.336 | 0.737 | 0.791 | 0.005 |
|  | SymbolSub | -0.029 | 0.015 | -1.893 | 0.058 | 0.135 | -0.062 |
|  | DigitSpan | 0.012 | 0.027 | 0.459 | 0.647 | 0.700 | 0.024 |
|  | TrailA | -0.025 | 0.019 | -1.323 | 0.186 | 0.327 | -0.047 |
|  | TrailB | -0.020 | 0.018 | -1.090 | 0.276 | 0.409 | -0.038 |
| N Childbirth | PairsMatch | 0.026 | 0.006 | 4.423 | **9.74e-06** | **4.43e-05** | 0.021 |
|  | ReactionTime | 0.064 | 0.006 | 11.277 | **1.75e-29** | **1.15e-27** | 0.054 |
|  | SymbolSub | -0.023 | 0.010 | -2.243 | **0.025** | 0.064 | -0.020 |
|  | DigitSpan | -0.013 | 0.018 | -0.713 | 0.476 | 0.582 | -0.011 |
|  | TrailA | -0.040 | 0.012 | -3.397 | **0.001** | **0.003** | -0.033 |
|  | TrailB | 0.007 | 0.011 | 0.615 | 0.538 | 0.623 | 0.006 |
| N Childbirth^2^ | PairsMatch | -0.012 | 0.006 | -1.955 | 0.051 | 0.119 | -0.009 |
|  | ReactionTime | -0.063 | 0.006 | -11.023 | **3.04e-28** | **1.34e-26** | -0.053 |
|  | SymbolSub | -0.017 | 0.011 | -1.502 | 0.133 | 0.252 | -0.014 |
|  | DigitSpan | -0.005 | 0.018 | -0.252 | 0.801 | 0.852 | -0.004 |
|  | TrailA | -0.014 | 0.013 | -1.070 | 0.285 | 0.413 | -0.010 |
|  | TrailB | -0.025 | 0.012 | -1.990 | **0.047** | 0.112 | -0.019 |
| Age last Birth | PairsMatch | 0.012 | 0.003 | 4.156 | **3.24e-05** | **1.38e-04** | 0.024 |
|  | ReactionTime | -0.003 | 0.003 | -0.872 | 0.383 | 0.521 | -0.005 |
|  | SymbolSub | 0.002 | 0.005 | 0.458 | 0.647 | 0.700 | 0.005 |
|  | DigitSpan | 0.022 | 0.009 | 2.451 | **0.014** | **0.038** | 0.044 |
|  | TrailA | 0.016 | 0.006 | 2.606 | **0.009** | **0.028** | 0.032 |
|  | TrailB | 0.016 | 0.006 | 2.753 | **0.006** | **0.019** | 0.034 |
| Age first Birth | PairsMatch | 0.022 | 0.003 | 7.191 | **6.48e-13** | **6.58e-12** | 0.042 |
|  | ReactionTime | 0.018 | 0.003 | 6.070 | **1.28e-09** | **9.41e-09** | 0.036 |
|  | SymbolSub | 0.032 | 0.005 | 5.982 | **2.23e-09** | **1.47e-08** | 0.068 |
|  | DigitSpan | 0.049 | 0.010 | 5.204 | **1.99e-07** | **1.05e-06** | 0.094 |
|  | TrailA | 0.043 | 0.006 | 6.914 | **4.83e-12** | **4.56e-11** | 0.085 |
|  | TrailB | 0.046 | 0.006 | 7.744 | **9.96e-15** | **1.31e-13** | 0.095 |
| Miscarriage & Termination | PairsMatch | -0.015 | 0.013 | -1.102 | 0.271 | 0.406 | -0.015 |
|  | ReactionTime | 0.008 | 0.013 | 0.631 | 0.528 | 0.623 | 0.009 |
|  | SymbolSub | 0.013 | 0.021 | 0.654 | 0.513 | 0.613 | 0.015 |
|  | DigitSpan | 0.029 | 0.042 | 0.684 | 0.494 | 0.598 | 0.030 |
|  | TrailA | 0.017 | 0.023 | 0.761 | 0.447 | 0.574 | 0.019 |
|  | TrailB | -0.012 | 0.022 | -0.553 | 0.580 | 0.650 | -0.014 |
| Stillbirth | PairsMatch | 0.004 | 0.077 | 0.058 | 0.954 | 0.968 | 0.005 |
|  | ReactionTime | -0.001 | 0.072 | -0.009 | 0.993 | 0.993 | -0.001 |
|  | SymbolSub | -0.018 | 0.131 | -0.138 | 0.890 | 0.918 | -0.021 |
|  | DigitSpan | -0.277 | 0.249 | -1.111 | 0.266 | 0.406 | -0.288 |
|  | TrailA | -0.254 | 0.152 | -1.663 | 0.096 | 0.199 | -0.274 |
|  | TrailB | -0.319 | 0.148 | -2.161 | **0.031** | 0.078 | -0.356 |
| Current HT use | PairsMatch | -0.031 | 0.011 | -2.911 | **0.004** | **0.013** | -0.026 |
|  | ReactionTime | 0.012 | 0.010 | 1.212 | 0.225 | 0.368 | 0.011 |
|  | SymbolSub | -0.013 | 0.017 | -0.729 | 0.466 | 0.580 | -0.012 |
|  | DigitSpan | 0.032 | 0.034 | 0.947 | 0.344 | 0.477 | 0.016 |
|  | TrailA | 0.040 | 0.020 | 1.993 | **0.046** | 0.112 | 0.022 |
|  | TrailB | -0.029 | 0.019 | -1.501 | 0.133 | 0.252 | -0.017 |
| Past HT use | PairsMatch | -0.033 | 0.006 | -5.394 | **6.89e-08** | **3.95e-07** | -0.048 |
|  | ReactionTime | 0.023 | 0.006 | 3.911 | **9.19e-05** | **3.68e-04** | 0.036 |
|  | SymbolSub | -0.029 | 0.011 | -2.763 | **0.006** | **0.019** | -0.046 |
|  | DigitSpan | 0.005 | 0.019 | 0.242 | 0.809 | 0.854 | 0.004 |
|  | TrailA | 0.010 | 0.013 | 0.826 | 0.409 | 0.550 | 0.009 |
|  | TrailB | -0.051 | 0.012 | -4.299 | **1.72e-05** | **7.56e-05** | -0.048 |
| Age HT initiation | PairsMatch | 0.000 | 0.005 | -0.088 | 0.930 | 0.951 | -0.001 |
|  | ReactionTime | 0.006 | 0.004 | 1.312 | 0.190 | 0.327 | 0.011 |
|  | SymbolSub | 0.012 | 0.008 | 1.500 | 0.134 | 0.252 | 0.025 |
|  | DigitSpan | 0.010 | 0.014 | 0.745 | 0.456 | 0.574 | 0.020 |
|  | TrailA | 0.005 | 0.010 | 0.552 | 0.581 | 0.650 | 0.010 |
|  | TrailB | 0.007 | 0.009 | 0.756 | 0.450 | 0.574 | 0.013 |
| Age HT rel. Menopause | PairsMatch | -0.003 | 0.005 | -0.586 | 0.558 | 0.635 | -0.006 |
|  | ReactionTime | -0.001 | 0.005 | -0.225 | 0.822 | 0.861 | -0.002 |
|  | SymbolSub | 0.007 | 0.009 | 0.791 | 0.429 | 0.572 | 0.016 |
|  | DigitSpan | -0.003 | 0.016 | -0.197 | 0.844 | 0.877 | -0.006 |
|  | TrailA | 0.012 | 0.011 | 1.041 | 0.298 | 0.423 | 0.022 |
|  | TrailB | 0.027 | 0.011 | 2.501 | **0.012** | **0.035** | 0.053 |
| Duration HT use | PairsMatch | -0.005 | 0.005 | -1.190 | 0.234 | 0.377 | -0.010 |
|  | ReactionTime | 0.005 | 0.005 | 1.109 | 0.267 | 0.406 | 0.009 |
|  | SymbolSub | -0.012 | 0.009 | -1.404 | 0.160 | 0.294 | -0.023 |
|  | DigitSpan | 0.010 | 0.014 | 0.744 | 0.457 | 0.574 | 0.020 |
|  | TrailA | 0.006 | 0.010 | 0.595 | 0.552 | 0.633 | 0.011 |
|  | TrailB | -0.016 | 0.010 | -1.644 | 0.100 | 0.203 | -0.030 |
| Current HC use | PairsMatch | 0.046 | 0.018 | 2.558 | **0.011** | **0.032** | 0.021 |
|  | ReactionTime | 0.145 | 0.017 | 8.440 | **3.19e-17** | **6.01e-16** | 0.059 |
|  | SymbolSub | 0.154 | 0.029 | 5.387 | **7.19e-08** | **3.95e-07** | 0.069 |
|  | DigitSpan | 0.198 | 0.054 | 3.666 | **2.47e-04** | **0.001** | 0.072 |
|  | TrailA | 0.158 | 0.033 | 4.836 | **1.33e-06** | **6.28e-06** | 0.068 |
|  | TrailB | 0.190 | 0.031 | 6.085 | **1.18e-09** | **9.15e-09** | 0.087 |
| Past HC use | PairsMatch | 0.016 | 0.006 | 2.521 | **0.012** | **0.034** | 0.020 |
|  | ReactionTime | 0.091 | 0.006 | 14.795 | **1.70e-49** | **2.25e-47** | 0.113 |
|  | SymbolSub | 0.105 | 0.012 | 9.023 | **1.89e-19** | **4.17e-18** | 0.116 |
|  | DigitSpan | 0.116 | 0.020 | 5.688 | **1.30e-08** | **7.83e-08** | 0.113 |
|  | TrailA | 0.094 | 0.014 | 6.763 | **1.37e-11** | **1.21e-10** | 0.089 |
|  | TrailB | 0.109 | 0.013 | 8.239 | **1.79e-16** | **2.95e-15** | 0.117 |
| Age HC initiation | PairsMatch | -0.018 | 0.003 | -5.970 | **2.37e-09** | **1.49e-08** | -0.032 |
|  | ReactionTime | -0.027 | 0.003 | -9.142 | **6.21e-20** | **1.64e-18** | -0.049 |
|  | SymbolSub | -0.050 | 0.005 | -9.676 | **4.04e-22** | **1.33e-20** | -0.096 |
|  | DigitSpan | -0.073 | 0.010 | -7.395 | **1.49e-13** | **1.79e-12** | -0.123 |
|  | TrailA | -0.030 | 0.006 | -4.961 | **7.04e-07** | **3.44e-06** | -0.053 |
|  | TrailB | -0.042 | 0.006 | -7.335 | **2.26e-13** | **2.49e-12** | -0.078 |
| Duration HC use | PairsMatch | 0.003 | 0.003 | 1.211 | 0.226 | 0.368 | 0.007 |
|  | ReactionTime | 0.005 | 0.003 | 1.786 | 0.074 | 0.166 | 0.010 |
|  | SymbolSub | 0.036 | 0.005 | 7.866 | **3.76e-15** | **5.51e-14** | 0.081 |
|  | DigitSpan | 0.044 | 0.009 | 5.168 | **2.40e-07** | **1.22e-06** | 0.089 |
|  | TrailA | 0.034 | 0.005 | 6.601 | **4.14e-11** | **3.42e-10** | 0.072 |
|  | TrailB | 0.030 | 0.005 | 6.057 | **1.40e-09** | **9.72e-09** | 0.066 |

Abbreviation: PairMatch = Pair Matching Test. ReactionTime = Reaction Time Test. SymbolSub = Symbol Substitution Test. DigitSpan = Digit Span Test. Trail A & B = Trail Making Test A & B. N = Number. HT = Hormone Therapy. HC = Hormonal Contraceptive. S.E. = Standard Error. FDR = False Discovery Rate. Significant results are highlighted in bold.

***Table S7| Associations between female-specific factors and late life cognition, including participants with ICD-10 diagnoses known to impact the brain and cognition.***

| **Model** | **Test** | **beta** | **S.E.** | **t** | **p** | **p_FDR_** | **d** |
| --- | --- | --- | --- | --- | --- | --- | --- |
| Reproductive Span | PairsMatch | 0.001 | 0.003 | 0.370 | 0.712 | 0.794 | 0.002 |
|  | ReactionTime | 0.014 | 0.003 | 4.368 | **1.25e-05** | **5.01e-05** | 0.026 |
|  | SymbolSub | 0.021 | 0.006 | 3.545 | **3.94e-04** | **0.001** | 0.042 |
|  | DigitSpan | 0.044 | 0.010 | 4.524 | **6.13e-06** | **2.53e-05** | 0.084 |
|  | TrailA | 0.011 | 0.007 | 1.552 | 0.121 | 0.213 | 0.020 |
|  | TrailB | 0.017 | 0.007 | 2.432 | **0.015** | **0.036** | 0.031 |
| Age at Menarche | PairsMatch | 0.003 | 0.002 | 1.279 | 0.201 | 0.320 | 0.006 |
|  | ReactionTime | -0.005 | 0.002 | -2.120 | **0.034** | 0.072 | -0.010 |
|  | SymbolSub | 0.005 | 0.004 | 1.236 | 0.216 | 0.332 | 0.011 |
|  | DigitSpan | 0.005 | 0.007 | 0.730 | 0.465 | 0.574 | 0.010 |
|  | TrailA | 0.013 | 0.005 | 2.761 | **0.006** | **0.014** | 0.026 |
|  | TrailB | 0.009 | 0.004 | 2.114 | **0.035** | 0.072 | 0.020 |
| Age at Menopause | PairsMatch | 0.003 | 0.003 | 0.928 | 0.353 | 0.471 | 0.005 |
|  | ReactionTime | 0.013 | 0.003 | 4.132 | **3.59e-05** | **1.40e-04** | 0.025 |
|  | SymbolSub | 0.023 | 0.006 | 3.778 | **1.59e-04** | **0.001** | 0.044 |
|  | DigitSpan | 0.048 | 0.010 | 4.938 | **8.00e-07** | **3.52e-06** | 0.091 |
|  | TrailA | 0.016 | 0.007 | 2.263 | **0.024** | 0.054 | 0.028 |
|  | TrailB | 0.022 | 0.007 | 3.148 | **0.002** | **0.005** | 0.040 |
| Oophorectomy | PairsMatch | 0.007 | 0.009 | 0.836 | 0.403 | 0.514 | 0.007 |
|  | ReactionTime | -0.018 | 0.008 | -2.183 | **0.029** | 0.065 | -0.018 |
|  | SymbolSub | 0.029 | 0.016 | 1.864 | 0.062 | 0.118 | 0.032 |
|  | DigitSpan | -0.052 | 0.026 | -2.000 | **0.046** | 0.090 | -0.051 |
|  | TrailA | 0.027 | 0.018 | 1.469 | 0.142 | 0.237 | 0.028 |
|  | TrailB | 0.014 | 0.018 | 0.769 | 0.442 | 0.550 | 0.014 |
| Age at Oophorectomy | PairsMatch | -0.014 | 0.009 | -1.525 | 0.127 | 0.221 | -0.025 |
|  | ReactionTime | 0.005 | 0.009 | 0.537 | 0.591 | 0.697 | 0.009 |
|  | SymbolSub | -0.005 | 0.018 | -0.274 | 0.784 | 0.835 | -0.009 |
|  | DigitSpan | 0.044 | 0.026 | 1.664 | 0.096 | 0.174 | 0.082 |
|  | TrailA | -0.027 | 0.022 | -1.263 | 0.207 | 0.325 | -0.046 |
|  | TrailB | -0.039 | 0.021 | -1.864 | 0.062 | 0.118 | -0.068 |
| Hysterectomy | PairsMatch | -0.001 | 0.008 | -0.151 | 0.880 | 0.915 | -0.001 |
|  | ReactionTime | 0.005 | 0.007 | 0.659 | 0.510 | 0.617 | 0.005 |
|  | SymbolSub | 0.015 | 0.014 | 1.079 | 0.281 | 0.399 | 0.017 |
|  | DigitSpan | -0.072 | 0.024 | -2.995 | **0.003** | **0.008** | -0.073 |
|  | TrailA | -0.014 | 0.017 | -0.833 | 0.405 | 0.514 | -0.014 |
|  | TrailB | 0.007 | 0.016 | 0.459 | 0.646 | 0.742 | 0.008 |
| Age at Hysterectomy | PairsMatch | -0.015 | 0.008 | -1.917 | 0.055 | 0.107 | -0.028 |
|  | ReactionTime | 0.008 | 0.007 | 1.153 | 0.249 | 0.361 | 0.017 |
|  | SymbolSub | -0.022 | 0.015 | -1.509 | 0.131 | 0.225 | -0.045 |
|  | DigitSpan | 0.008 | 0.024 | 0.348 | 0.728 | 0.800 | 0.016 |
|  | TrailA | -0.015 | 0.018 | -0.878 | 0.380 | 0.502 | -0.029 |
|  | TrailB | -0.004 | 0.017 | -0.210 | 0.834 | 0.881 | -0.007 |
| N Childbirth | PairsMatch | 0.023 | 0.004 | 5.607 | **2.07e-08** | **1.05e-07** | 0.025 |
|  | ReactionTime | 0.038 | 0.004 | 9.282 | **1.68e-20** | **4.44e-19** | 0.041 |
|  | SymbolSub | -0.027 | 0.009 | -3.006 | **0.003** | **0.008** | -0.026 |
|  | DigitSpan | -0.019 | 0.016 | -1.171 | 0.242 | 0.359 | -0.016 |
|  | TrailA | -0.043 | 0.011 | -4.119 | **3.81e-05** | **1.44e-04** | -0.038 |
|  | TrailB | 0.009 | 0.010 | 0.848 | 0.396 | 0.514 | 0.008 |
| N Childbirth^2^ | PairsMatch | -0.010 | 0.004 | -2.451 | **0.014** | **0.035** | -0.011 |
|  | ReactionTime | -0.037 | 0.004 | -9.037 | **1.62e-19** | **3.57e-18** | -0.040 |
|  | SymbolSub | -0.014 | 0.011 | -1.281 | 0.200 | 0.320 | -0.011 |
|  | DigitSpan | 0.001 | 0.018 | 0.068 | 0.946 | 0.960 | 0.001 |
|  | TrailA | -0.011 | 0.013 | -0.841 | 0.400 | 0.514 | -0.008 |
|  | TrailB | -0.030 | 0.012 | -2.367 | **0.018** | **0.042** | -0.022 |
| Age last Birth | PairsMatch | 0.009 | 0.003 | 3.084 | **0.002** | **0.006** | 0.017 |
|  | ReactionTime | -0.001 | 0.003 | -0.316 | 0.752 | 0.814 | -0.002 |
|  | SymbolSub | 0.002 | 0.005 | 0.429 | 0.668 | 0.753 | 0.005 |
|  | DigitSpan | 0.025 | 0.008 | 2.936 | **0.003** | **0.009** | 0.049 |
|  | TrailA | 0.017 | 0.006 | 2.906 | **0.004** | **0.010** | 0.034 |
|  | TrailB | 0.018 | 0.006 | 3.234 | **0.001** | **0.004** | 0.038 |
| Age first Birth | PairsMatch | 0.022 | 0.003 | 7.484 | **7.23e-14** | **6.36e-13** | 0.041 |
|  | ReactionTime | 0.022 | 0.003 | 8.052 | **8.20e-16** | **1.08e-14** | 0.044 |
|  | SymbolSub | 0.033 | 0.005 | 6.398 | **1.60e-10** | **1.24e-09** | 0.069 |
|  | DigitSpan | 0.050 | 0.009 | 5.734 | **1.00e-08** | **5.30e-08** | 0.096 |
|  | TrailA | 0.046 | 0.006 | 7.650 | **2.07e-14** | **1.96e-13** | 0.089 |
|  | TrailB | 0.049 | 0.006 | 8.622 | **6.90e-18** | **1.01e-16** | 0.101 |
| Miscarriage & Termination | PairsMatch | -0.014 | 0.013 | -1.137 | 0.256 | 0.367 | -0.014 |
|  | ReactionTime | 0.015 | 0.012 | 1.291 | 0.197 | 0.320 | 0.016 |
|  | SymbolSub | 0.012 | 0.019 | 0.639 | 0.523 | 0.627 | 0.014 |
|  | DigitSpan | 0.031 | 0.038 | 0.814 | 0.416 | 0.523 | 0.033 |
|  | TrailA | 0.008 | 0.022 | 0.364 | 0.716 | 0.794 | 0.009 |
|  | TrailB | -0.021 | 0.021 | -0.979 | 0.328 | 0.446 | -0.023 |
| Stillbirth | PairsMatch | -0.004 | 0.069 | -0.058 | 0.954 | 0.961 | -0.004 |
|  | ReactionTime | -0.012 | 0.066 | -0.175 | 0.861 | 0.902 | -0.012 |
|  | SymbolSub | -0.005 | 0.124 | -0.037 | 0.970 | 0.970 | -0.005 |
|  | DigitSpan | -0.216 | 0.226 | -0.953 | 0.341 | 0.459 | -0.226 |
|  | TrailA | -0.247 | 0.144 | -1.714 | 0.087 | 0.159 | -0.265 |
|  | TrailB | -0.399 | 0.139 | -2.862 | **0.004** | **0.011** | -0.443 |
| Current HT use | PairsMatch | -0.032 | 0.010 | -3.335 | **0.001** | **0.003** | -0.027 |
|  | ReactionTime | 0.013 | 0.009 | 1.404 | 0.160 | 0.265 | 0.011 |
|  | SymbolSub | -0.008 | 0.016 | -0.513 | 0.608 | 0.704 | -0.008 |
|  | DigitSpan | 0.038 | 0.031 | 1.238 | 0.216 | 0.332 | 0.019 |
|  | TrailA | 0.038 | 0.019 | 2.004 | **0.045** | 0.090 | 0.021 |
|  | TrailB | -0.030 | 0.018 | -1.656 | 0.098 | 0.174 | -0.017 |
| Past HT use | PairsMatch | -0.034 | 0.006 | -5.885 | **4.00e-09** | **2.29e-08** | -0.047 |
|  | ReactionTime | 0.020 | 0.005 | 3.707 | **2.10e-04** | **0.001** | 0.031 |
|  | SymbolSub | -0.032 | 0.010 | -3.166 | **0.002** | **0.005** | -0.049 |
|  | DigitSpan | 0.005 | 0.018 | 0.303 | 0.762 | 0.817 | 0.005 |
|  | TrailA | 0.012 | 0.012 | 0.995 | 0.320 | 0.443 | 0.010 |
|  | TrailB | -0.051 | 0.011 | -4.534 | **5.79e-06** | **2.47e-05** | -0.048 |
| Age HT initiation | PairsMatch | -4.23e-04 | 0.004 | -0.101 | 0.920 | 0.941 | -0.001 |
|  | ReactionTime | 0.009 | 0.004 | 2.129 | **0.033** | 0.072 | 0.016 |
|  | SymbolSub | 0.021 | 0.008 | 2.817 | **0.005** | **0.013** | 0.043 |
|  | DigitSpan | 0.028 | 0.013 | 2.163 | **0.031** | 0.067 | 0.052 |
|  | TrailA | 0.004 | 0.009 | 0.445 | 0.656 | 0.746 | 0.007 |
|  | TrailB | 0.016 | 0.009 | 1.764 | 0.078 | 0.145 | 0.029 |
| Age HT rel. Menopause | PairsMatch | -0.001 | 0.005 | -0.134 | 0.893 | 0.921 | -0.001 |
|  | ReactionTime | -0.003 | 0.005 | -0.691 | 0.489 | 0.598 | -0.006 |
|  | SymbolSub | 0.005 | 0.009 | 0.520 | 0.603 | 0.704 | 0.009 |
|  | DigitSpan | -0.016 | 0.015 | -1.039 | 0.299 | 0.419 | -0.030 |
|  | TrailA | 0.004 | 0.011 | 0.339 | 0.735 | 0.801 | 0.007 |
|  | TrailB | 0.021 | 0.011 | 2.018 | **0.044** | 0.089 | 0.040 |
| Duration HT use | PairsMatch | -0.004 | 0.004 | -0.990 | 0.322 | 0.443 | -0.008 |
|  | ReactionTime | 0.005 | 0.004 | 1.152 | 0.249 | 0.361 | 0.009 |
|  | SymbolSub | -0.009 | 0.008 | -1.175 | 0.240 | 0.359 | -0.018 |
|  | DigitSpan | 0.007 | 0.013 | 0.590 | 0.555 | 0.660 | 0.014 |
|  | TrailA | 0.011 | 0.009 | 1.193 | 0.233 | 0.353 | 0.020 |
|  | TrailB | -0.018 | 0.009 | -2.046 | **0.041** | 0.084 | -0.034 |
| Current HC use | PairsMatch | 0.047 | 0.017 | 2.769 | **0.006** | **0.014** | 0.021 |
|  | ReactionTime | 0.152 | 0.016 | 9.328 | **1.09e-20** | **3.59e-19** | 0.062 |
|  | SymbolSub | 0.150 | 0.028 | 5.462 | **4.73e-08** | **2.23e-07** | 0.067 |
|  | DigitSpan | 0.185 | 0.051 | 3.628 | **2.86e-04** | **0.001** | 0.066 |
|  | TrailA | 0.158 | 0.031 | 5.053 | **4.36e-07** | **1.98e-06** | 0.067 |
|  | TrailB | 0.173 | 0.030 | 5.775 | **7.74e-09** | **4.25e-08** | 0.078 |
| Past HC use | PairsMatch | 0.014 | 0.006 | 2.349 | **0.019** | **0.044** | 0.018 |
|  | ReactionTime | 0.092 | 0.006 | 16.023 | **9.64e-58** | **1.27e-55** | 0.116 |
|  | SymbolSub | 0.100 | 0.011 | 9.009 | **2.16e-19** | **4.07e-18** | 0.111 |
|  | DigitSpan | 0.111 | 0.019 | 5.887 | **3.99e-09** | **2.29e-08** | 0.107 |
|  | TrailA | 0.083 | 0.013 | 6.338 | **2.35e-10** | **1.73e-09** | 0.079 |
|  | TrailB | 0.098 | 0.013 | 7.764 | **8.39e-15** | **8.52e-14** | 0.105 |
| Age HC initiation | PairsMatch | -0.017 | 0.003 | -6.007 | **1.89e-09** | **1.19e-08** | -0.030 |
|  | ReactionTime | -0.028 | 0.003 | -10.586 | **3.51e-26** | **1.55e-24** | -0.053 |
|  | SymbolSub | -0.052 | 0.005 | -10.966 | **6.07e-28** | **4.01e-26** | -0.103 |
|  | DigitSpan | -0.062 | 0.009 | -7.153 | **8.82e-13** | **7.28e-12** | -0.109 |
|  | TrailA | -0.035 | 0.006 | -6.217 | **5.11e-10** | **3.37e-09** | -0.063 |
|  | TrailB | -0.042 | 0.005 | -7.841 | **4.60e-15** | **5.52e-14** | -0.079 |
| Duration HC use | PairsMatch | 0.004 | 0.003 | 1.470 | 0.141 | 0.237 | 0.008 |
|  | ReactionTime | 0.008 | 0.002 | 3.424 | **0.001** | **0.002** | 0.018 |
|  | SymbolSub | 0.038 | 0.004 | 8.900 | **5.81e-19** | **9.59e-18** | 0.086 |
|  | DigitSpan | 0.043 | 0.008 | 5.482 | **4.26e-08** | **2.08e-07** | 0.087 |
|  | TrailA | 0.038 | 0.005 | 7.803 | **6.18e-15** | **6.80e-14** | 0.081 |
|  | TrailB | 0.029 | 0.005 | 6.227 | **4.80e-10** | **3.33e-09** | 0.064 |

Abbreviation: PairMatch = Pair Matching Test. ReactionTime = Reaction Time Test. SymbolSub = Symbol Substitution Test. DigitSpan = Digit Span Test. Trail A & B = Trail Making Test A & B. N = Number. HT = Hormone Therapy. HC = Hormonal Contraceptive. S.E. = Standard Error. FDR = False Discovery Rate. Significant results are highlighted in bold.

***Table S8| Associations between female-specific factors and late life cognition after adjustment for APOE ε4*** ***genotype.***

| **Model** | **Test** | **beta** | **S.E.** | **t** | **p** | **p_FDR_** | **d** |
| --- | --- | --- | --- | --- | --- | --- | --- |
| Reproductive Span | PairsMatch | 0.002 | 0.004 | 0.662 | 0.508 | 0.612 | 0.004 |
|  | ReactionTime | 0.008 | 0.003 | 2.444 | **0.015** | **0.038** | 0.016 |
|  | SymbolSub | 0.015 | 0.006 | 2.414 | **0.016** | **0.041** | 0.030 |
|  | DigitSpan | 0.037 | 0.011 | 3.404 | **0.001** | **0.002** | 0.070 |
|  | TrailA | 0.006 | 0.008 | 0.836 | 0.403 | 0.537 | 0.011 |
|  | TrailB | 0.008 | 0.007 | 1.080 | 0.280 | 0.425 | 0.015 |
| Age at Menarche | PairsMatch | 0.002 | 0.002 | 0.934 | 0.350 | 0.503 | 0.005 |
|  | ReactionTime | -0.004 | 0.002 | -1.680 | 0.093 | 0.195 | -0.008 |
|  | SymbolSub | 0.006 | 0.004 | 1.335 | 0.182 | 0.324 | 0.013 |
|  | DigitSpan | 0.004 | 0.008 | 0.524 | 0.600 | 0.666 | 0.008 |
|  | TrailA | 0.013 | 0.005 | 2.521 | **0.012** | **0.032** | 0.025 |
|  | TrailB | 0.007 | 0.005 | 1.490 | 0.136 | 0.264 | 0.015 |
| Age at Menopause | PairsMatch | 0.004 | 0.004 | 1.072 | 0.284 | 0.426 | 0.007 |
|  | ReactionTime | 0.007 | 0.003 | 2.029 | **0.043** | 0.098 | 0.013 |
|  | SymbolSub | 0.017 | 0.006 | 2.640 | **0.008** | **0.024** | 0.033 |
|  | DigitSpan | 0.040 | 0.011 | 3.744 | **1.82e-04** | **0.001** | 0.076 |
|  | TrailA | 0.011 | 0.008 | 1.398 | 0.162 | 0.301 | 0.019 |
|  | TrailB | 0.011 | 0.007 | 1.468 | 0.142 | 0.272 | 0.020 |
| Oophorectomy | PairsMatch | 0.011 | 0.010 | 1.147 | 0.251 | 0.400 | 0.011 |
|  | ReactionTime | -0.021 | 0.009 | -2.301 | **0.021** | 0.051 | -0.022 |
|  | SymbolSub | 0.041 | 0.017 | 2.369 | **0.018** | **0.045** | 0.045 |
|  | DigitSpan | -0.039 | 0.030 | -1.328 | 0.184 | 0.324 | -0.039 |
|  | TrailA | 0.031 | 0.020 | 1.498 | 0.134 | 0.264 | 0.031 |
|  | TrailB | 0.017 | 0.020 | 0.881 | 0.378 | 0.515 | 0.018 |
| Age at Oophorectomy | PairsMatch | -0.017 | 0.010 | -1.638 | 0.101 | 0.209 | -0.030 |
|  | ReactionTime | -0.006 | 0.010 | -0.624 | 0.533 | 0.622 | -0.012 |
|  | SymbolSub | -0.005 | 0.020 | -0.278 | 0.781 | 0.825 | -0.010 |
|  | DigitSpan | 0.038 | 0.030 | 1.270 | 0.204 | 0.339 | 0.072 |
|  | TrailA | -0.030 | 0.024 | -1.266 | 0.206 | 0.339 | -0.051 |
|  | TrailB | -0.049 | 0.023 | -2.128 | **0.033** | 0.079 | -0.087 |
| Hysterectomy | PairsMatch | -0.004 | 0.009 | -0.487 | 0.626 | 0.683 | -0.002 |
|  | ReactionTime | 0.003 | 0.008 | 0.384 | 0.701 | 0.752 | 0.003 |
|  | SymbolSub | 0.020 | 0.015 | 1.290 | 0.197 | 0.339 | 0.022 |
|  | DigitSpan | -0.071 | 0.028 | -2.571 | **0.010** | **0.029** | -0.071 |
|  | TrailA | 0.002 | 0.018 | 0.130 | 0.897 | 0.932 | 0.002 |
|  | TrailB | 0.026 | 0.017 | 1.512 | 0.130 | 0.264 | 0.028 |
| Age at Hysterectomy | PairsMatch | -0.011 | 0.009 | -1.230 | 0.219 | 0.357 | -0.020 |
|  | ReactionTime | 0.006 | 0.008 | 0.674 | 0.501 | 0.612 | 0.011 |
|  | SymbolSub | -0.031 | 0.016 | -1.959 | **0.050** | 0.110 | -0.065 |
|  | DigitSpan | 0.002 | 0.027 | 0.087 | 0.931 | 0.952 | 0.005 |
|  | TrailA | -0.022 | 0.019 | -1.132 | 0.258 | 0.404 | -0.041 |
|  | TrailB | -0.017 | 0.018 | -0.909 | 0.364 | 0.511 | -0.033 |
| N Childbirth | PairsMatch | 0.025 | 0.004 | 5.701 | **1.19e-08** | **7.73e-08** | 0.028 |
|  | ReactionTime | 0.032 | 0.004 | 7.434 | **1.06e-13** | **1.55e-12** | 0.036 |
|  | SymbolSub | -0.031 | 0.010 | -3.052 | **0.002** | **0.008** | -0.028 |
|  | DigitSpan | -0.009 | 0.018 | -0.528 | 0.598 | 0.666 | -0.008 |
|  | TrailA | -0.046 | 0.012 | -3.975 | **7.06e-05** | **2.91e-04** | -0.040 |
|  | TrailB | 0.008 | 0.011 | 0.711 | 0.477 | 0.597 | 0.007 |
| N Childbirth^2^ | PairsMatch | -0.012 | 0.004 | -2.712 | **0.007** | **0.021** | -0.013 |
|  | ReactionTime | -0.031 | 0.004 | -7.098 | **1.27e-12** | 1.19e-11 | -0.035 |
|  | SymbolSub | -0.011 | 0.013 | -0.864 | 0.387 | 0.522 | -0.008 |
|  | DigitSpan | -0.010 | 0.020 | -0.530 | 0.596 | 0.666 | -0.008 |
|  | TrailA | -0.011 | 0.015 | -0.725 | 0.468 | 0.595 | -0.007 |
|  | TrailB | -0.033 | 0.014 | -2.324 | **0.020** | **0.049** | -0.023 |
| Age last Birth | PairsMatch | 0.011 | 0.003 | 3.444 | **0.001** | **0.002** | 0.021 |
|  | ReactionTime | -0.002 | 0.003 | -0.796 | 0.426 | 0.557 | -0.005 |
|  | SymbolSub | 0.000 | 0.005 | 0.069 | 0.945 | 0.960 | 0.001 |
|  | DigitSpan | 0.027 | 0.009 | 2.916 | **0.004** | **0.012** | 0.054 |
|  | TrailA | 0.016 | 0.006 | 2.514 | **0.012** | **0.032** | 0.032 |
|  | TrailB | 0.012 | 0.006 | 2.000 | **0.046** | 0.102 | 0.025 |
| Age first Birth | PairsMatch | 0.023 | 0.003 | 7.173 | **7.39e-13** | **8.06e-12** | 0.043 |
|  | ReactionTime | 0.020 | 0.003 | 6.545 | **5.97e-11** | **4.92e-10** | 0.039 |
|  | SymbolSub | 0.031 | 0.005 | 5.689 | **1.29e-08** | **7.73e-08** | 0.066 |
|  | DigitSpan | 0.051 | 0.010 | 5.264 | **1.43e-07** | **7.56e-07** | 0.097 |
|  | TrailA | 0.046 | 0.006 | 7.166 | **7.93e-13** | **8.06e-12** | 0.090 |
|  | TrailB | 0.044 | 0.006 | 7.247 | **4.37e-13** | **5.24e-12** | 0.091 |
| Miscarriage & Termination | PairsMatch | -0.020 | 0.014 | -1.411 | 0.158 | 0.299 | -0.020 |
|  | ReactionTime | 0.012 | 0.013 | 0.888 | 0.375 | 0.515 | 0.012 |
|  | SymbolSub | 0.015 | 0.021 | 0.707 | 0.480 | 0.597 | 0.017 |
|  | DigitSpan | 0.028 | 0.043 | 0.639 | 0.523 | 0.616 | 0.029 |
|  | TrailA | 0.014 | 0.023 | 0.594 | 0.552 | 0.634 | 0.015 |
|  | TrailB | -0.015 | 0.023 | -0.652 | 0.515 | 0.612 | -0.017 |
| Stillbirth | PairsMatch | -0.001 | 0.080 | -0.018 | 0.986 | 0.986 | -0.001 |
|  | ReactionTime | 0.037 | 0.075 | 0.494 | 0.621 | 0.683 | 0.040 |
|  | SymbolSub | -0.004 | 0.135 | -0.030 | 0.976 | 0.984 | -0.005 |
|  | DigitSpan | -0.169 | 0.259 | -0.654 | 0.513 | 0.612 | -0.176 |
|  | TrailA | -0.274 | 0.157 | -1.747 | 0.081 | 0.172 | -0.296 |
|  | TrailB | -0.280 | 0.152 | -1.839 | 0.066 | 0.143 | -0.312 |
| Current HT use | PairsMatch | -0.031 | 0.011 | -2.861 | **0.004** | **0.014** | -0.026 |
|  | ReactionTime | 0.012 | 0.010 | 1.126 | 0.260 | 0.404 | 0.010 |
|  | SymbolSub | -0.010 | 0.018 | -0.555 | 0.579 | 0.659 | -0.009 |
|  | DigitSpan | 0.032 | 0.035 | 0.927 | 0.354 | 0.503 | 0.016 |
|  | TrailA | 0.041 | 0.021 | 2.008 | **0.045** | 0.102 | 0.023 |
|  | TrailB | -0.025 | 0.020 | -1.288 | 0.198 | 0.339 | -0.015 |
| Past HT use | PairsMatch | -0.033 | 0.006 | -5.152 | **2.58e-07** | **1.26e-06** | -0.047 |
|  | ReactionTime | 0.025 | 0.006 | 4.170 | **3.05e-05** | **1.30e-04** | 0.039 |
|  | SymbolSub | -0.030 | 0.011 | -2.777 | **0.005** | **0.018** | -0.047 |
|  | DigitSpan | 0.007 | 0.020 | 0.353 | 0.724 | 0.771 | 0.006 |
|  | TrailA | 0.008 | 0.013 | 0.615 | 0.538 | 0.623 | 0.007 |
|  | TrailB | -0.053 | 0.012 | -4.324 | **1.53e-05** | **6.75e-05** | -0.049 |
| Age HT initiation | PairsMatch | -0.003 | 0.005 | -0.737 | 0.461 | 0.591 | -0.006 |
|  | ReactionTime | 0.005 | 0.005 | 1.188 | 0.235 | 0.378 | 0.010 |
|  | SymbolSub | 0.019 | 0.008 | 2.365 | **0.018** | **0.045** | 0.040 |
|  | DigitSpan | 0.015 | 0.014 | 1.041 | 0.298 | 0.442 | 0.028 |
|  | TrailA | 0.001 | 0.010 | 0.095 | 0.924 | 0.952 | 0.002 |
|  | TrailB | 0.007 | 0.010 | 0.765 | 0.444 | 0.575 | 0.014 |
| Age HT rel. Menopause | PairsMatch | -0.004 | 0.005 | -0.816 | 0.414 | 0.547 | -0.008 |
|  | ReactionTime | 0.001 | 0.005 | 0.187 | 0.852 | 0.892 | 0.002 |
|  | SymbolSub | 0.010 | 0.010 | 1.081 | 0.280 | 0.425 | 0.021 |
|  | DigitSpan | -0.007 | 0.017 | -0.427 | 0.669 | 0.724 | -0.014 |
|  | TrailA | 0.011 | 0.012 | 0.965 | 0.335 | 0.485 | 0.021 |
|  | TrailB | 0.030 | 0.011 | 2.574 | **0.010** | **0.029** | 0.055 |
| Duration HT use | PairsMatch | -0.004 | 0.005 | -0.884 | 0.377 | 0.515 | -0.008 |
|  | ReactionTime | 0.004 | 0.005 | 0.981 | 0.326 | 0.479 | 0.009 |
|  | SymbolSub | -0.013 | 0.009 | -1.505 | 0.132 | 0.264 | -0.025 |
|  | DigitSpan | 0.009 | 0.014 | 0.666 | 0.506 | 0.612 | 0.018 |
|  | TrailA | 0.013 | 0.010 | 1.280 | 0.201 | 0.339 | 0.023 |
|  | TrailB | -0.013 | 0.010 | -1.383 | 0.167 | 0.306 | -0.025 |
| Current HC use | PairsMatch | 0.050 | 0.018 | 2.692 | **0.007** | **0.022** | 0.022 |
|  | ReactionTime | 0.135 | 0.018 | 7.696 | **1.41e-14** | **3.10e-13** | 0.055 |
|  | SymbolSub | 0.151 | 0.029 | 5.161 | **2.46e-07** | **1.25e-06** | 0.068 |
|  | DigitSpan | 0.203 | 0.056 | 3.645 | **2.68e-04** | **0.001** | 0.074 |
|  | TrailA | 0.162 | 0.033 | 4.866 | **1.15e-06** | **5.21e-06** | 0.070 |
|  | TrailB | 0.195 | 0.032 | 6.134 | **8.64e-10** | **6.34e-09** | 0.089 |
| Past HC use | PairsMatch | 0.018 | 0.007 | 2.650 | **0.008** | **0.024** | 0.022 |
|  | ReactionTime | 0.092 | 0.006 | 14.549 | **6.34e-48** | **8.37e-46** | 0.114 |
|  | SymbolSub | 0.103 | 0.012 | 8.586 | **9.30e-18** | **3.07e-16** | 0.112 |
|  | DigitSpan | 0.119 | 0.021 | 5.695 | **1.26e-08** | **7.73e-08** | 0.116 |
|  | TrailA | 0.087 | 0.014 | 6.159 | **7.40e-10** | **5.75e-09** | 0.083 |
|  | TrailB | 0.103 | 0.014 | 7.591 | **3.25e-14** | **5.36e-13** | 0.111 |
| Age HC initiation | PairsMatch | -0.016 | 0.003 | -5.113 | **3.17e-07** | **1.50e-06** | -0.028 |
|  | ReactionTime | -0.028 | 0.003 | -9.809 | **1.04e-22** | **4.59e-21** | -0.053 |
|  | SymbolSub | -0.050 | 0.005 | -9.878 | **5.52e-23** | **3.64e-21** | -0.100 |
|  | DigitSpan | -0.071 | 0.010 | -7.396 | **1.48e-13** | **1.96e-12** | -0.125 |
|  | TrailA | -0.032 | 0.006 | -5.334 | **9.68e-08** | **5.33e-07** | -0.058 |
|  | TrailB | -0.043 | 0.006 | -7.603 | **2.98e-14** | **5.36e-13** | -0.082 |
| Duration HC use | PairsMatch | 0.004 | 0.003 | 1.361 | 0.173 | 0.314 | 0.008 |
|  | ReactionTime | 0.008 | 0.003 | 2.826 | **0.005** | **0.016** | 0.016 |
|  | SymbolSub | 0.036 | 0.005 | 7.877 | **3.45e-15** | **9.11e-14** | 0.082 |
|  | DigitSpan | 0.049 | 0.009 | 5.670 | **1.46e-08** | **8.39e-08** | 0.099 |
|  | TrailA | 0.035 | 0.005 | 6.745 | **1.55e-11** | **1.37e-10** | 0.075 |
|  | TrailB | 0.029 | 0.005 | 5.745 | **9.25e-09** | **6.43e-08** | 0.064 |

Abbreviation: APOE = Apolipoprotein, PairMatch = Pair Matching Test, ReactionTime = Reaction Time Test, SymbolSub = Symbol Substitution Test, DigitSpan = Digit Span Test, Trail A & B = Trail Making Test A & B, N = Number, HT = Hormone Therapy, HC = Hormonal Contraceptive, S.E. = Standard Error, FDR = False Discovery Rate. Significant results are highlighted in bold.

***Table S9****|* ***Detected extreme values of continuous female-specific factors using the median absolute deviation method.***

|  |  |  |  | **Number extreme values** | | |
| --- | --- | --- | --- | --- | --- | --- |
| **Variable** | **Median** | **MAD** | **Limits*** | **Low** | **High** | **Total** |
| Age at Menarche | 13 | 1.48 | 8.55 – 17.45 | 130 | 920 | 1050 |
| Age at Menopause | 50 | 4.45 | 36.66 – 63.34 | 2200 | 24 | 2224 |
| Age at Bilateral Oophorectomy | 48 | 7.41 | 25.76 – 70.24 | 51 | 0 | 51 |
| Age at Hysterectomy | 44 | 7.41 | 21.76 – 66.24 | 26 | 140 | 166 |
| Number of live Childbirths | 2 | 1.48 | -2.45 – 6.45 | 0 | 322 | 322 |
| Age at first Childbirth | 25 | 4.45 | 11.66 – 38.34 | 1 | 790 | 791 |
| Age at last Childbirth | 30 | 4.45 | 16.66 – 43.34 | 7 | 497 | 504 |
| Age started HT | 48 | 4.45 | 34.66 – 61.34 | 1185 | 115 | 1300 |
| Age last used HT | 55 | 5.93 | 37.21 – 72.79 | 537 | 0 | 537 |
| Age started HC | 21 | 4.45 | 7.66 – 34.34 | 10 | 3364 | 3374 |
| Age last used HC | 31 | 7.41 | 8.76 – 53.24 | 2 | 741 | 743 |

*Limits of acceptable range of values. Abbreviations: HT = hormone therapy, HC = hormonal contraceptives, MAD = median absolute deviation.

***Table S10| Main effects of APOE ε4*** ***genotype on cognitive performance.***

| **Model** | **Test** | **beta** | **S.E.** | **t** | **p** | **pFDR** | **d** |
| --- | --- | --- | --- | --- | --- | --- | --- |
| APOE ε4 | PairsMatch | -0.010 | 0.005 | -1.873 | 0.061 | 0.123 | -0.010 |
|  | ReactionTime | -0.001 | 0.005 | -0.162 | 0.871 | 0.925 | -0.001 |
|  | SymbolSub | -0.046 | 0.009 | -4.974 | **6.59e-07** | **5.93e-06** | -0.053 |
|  | DigitSpan | -0.031 | 0.017 | -1.823 | 0.068 | 0.123 | -0.032 |
|  | TrailA | -0.014 | 0.011 | -1.267 | 0.205 | 0.284 | -0.014 |
|  | TrailB | -0.027 | 0.010 | -2.673 | **0.008** | **0.027** | -0.030 |
| 1x ε4 allele | PairsMatch | -0.008 | 0.006 | -1.417 | 0.156 | 0.242 | -0.008 |
| 2x ε4 allele | PairsMatch | -0.033 | 0.016 | -2.060 | **0.039** | 0.118 | -0.012 |
| 1x ε4 allele | ReactionTime | -0.001 | 0.005 | -0.145 | 0.884 | 0.925 | -0.001 |
| 2x ε4 allele | ReactionTime | -0.001 | 0.015 | -0.094 | 0.925 | 0.925 | -0.001 |
| 1x ε4 allele | SymbolSub | -0.037 | 0.010 | -3.840 | **1.23e-04** | **0.001** | -0.042 |
| 2x ε4 allele | SymbolSub | -0.145 | 0.027 | -5.276 | **1.33e-07** | **2.39e-06** | -0.057 |
| 1x ε4 allele | DigitSpan | -0.034 | 0.018 | -1.964 | 0.049 | 0.123 | -0.086 |
| 2x ε4 allele | DigitSpan | 0.005 | 0.049 | 0.109 | 0.913 | 0.925 | 0.005 |
| 1x ε4 allele | TrailA | -0.011 | 0.011 | -0.962 | 0.336 | 0.432 | -0.028 |
| 2x ε4 allele | TrailA | -0.045 | 0.032 | -1.401 | 0.161 | 0.242 | -0.040 |
| 1x ε4 allele | TrailB | -0.020 | 0.011 | -1.852 | 0.064 | 0.123 | -0.055 |
| 2x ε4 allele | TrailB | -0.111 | 0.031 | -3.619 | **2.96e-04** | **0.001** | -0.107 |

Abbreviation: APOE = Apolipoprotein, PairMatch = Pair Matching Test, ReactionTime = Reaction Time Test, SymbolSub = Symbol Substitution Test, DigitSpan = Digit Span Test, Trail A & B = Trail Making Test A & B, N = Number, HT = Hormone Therapy, HC = Hormonal Contraceptive, S.E. = Standard Error, FDR = False Discovery Rate. Significant results are highlighted in bold.

***Table 11| Interactions between APOE ε4 genotype and female-specific factors on late life cognition.***

| **Model** | **Test** | **beta** | **S.E.** | **t** | **p** | **p_FDR_** |
| --- | --- | --- | --- | --- | --- | --- |
| Reproductive Span * *APOE* ε4 | PairsMatch | -0.002 | 0.002 | -0.783 | 0.434 | 0.867 |
|  | ReactionTime | 0.001 | 0.002 | 0.494 | 0.621 | 0.867 |
|  | SymbolSub | 0.006 | 0.004 | 1.353 | 0.176 | 0.867 |
|  | DigitSpan | 0.007 | 0.007 | 0.987 | 0.324 | 0.867 |
|  | TrailA | -0.007 | 0.005 | -1.359 | 0.174 | 0.867 |
|  | TrailB | 0.003 | 0.005 | 0.671 | 0.502 | 0.867 |
| Age at Menarche * *APOE* ε4 | PairsMatch | 0.003 | 0.007 | 0.514 | 0.607 | 0.867 |
|  | ReactionTime | -0.005 | 0.006 | -0.752 | 0.452 | 0.867 |
|  | SymbolSub | -0.001 | 0.012 | -0.047 | 0.963 | 0.983 |
|  | DigitSpan | 0.008 | 0.020 | 0.394 | 0.693 | 0.897 |
|  | TrailA | 0.003 | 0.014 | 0.218 | 0.828 | 0.966 |
|  | TrailB | 0.007 | 0.013 | 0.499 | 0.617 | 0.867 |
| Age at Menopause * *APOE* ε4 | PairsMatch | -0.004 | 0.007 | -0.598 | 0.550 | 0.867 |
|  | ReactionTime | 0.004 | 0.006 | 0.569 | 0.569 | 0.867 |
|  | SymbolSub | -0.008 | 0.012 | -0.706 | 0.480 | 0.867 |
|  | DigitSpan | 0.019 | 0.021 | 0.929 | 0.353 | 0.867 |
|  | TrailA | -0.002 | 0.014 | -0.160 | 0.873 | 0.968 |
|  | TrailB | 0.011 | 0.013 | 0.826 | 0.409 | 0.867 |
| Oophorectomy * *APOE* ε4 | PairsMatch | -0.002 | 0.014 | -0.119 | 0.906 | 0.977 |
|  | ReactionTime | -0.029 | 0.013 | -2.210 | 0.027 | 0.596 |
|  | SymbolSub | 0.023 | 0.021 | 1.098 | 0.272 | 0.867 |
|  | DigitSpan | -0.028 | 0.043 | -0.653 | 0.514 | 0.867 |
|  | TrailA | 0.031 | 0.024 | 1.322 | 0.186 | 0.867 |
|  | TrailB | 0.013 | 0.023 | 0.553 | 0.580 | 0.867 |
| Age at Oophorectomy * *APOE* ε4 | PairsMatch | -0.003 | 0.003 | -1.033 | 0.301 | 0.867 |
|  | ReactionTime | 0.005 | 0.003 | 1.997 | 0.046 | 0.755 |
|  | SymbolSub | -0.001 | 0.005 | -0.220 | 0.826 | 0.966 |
|  | DigitSpan | -0.008 | 0.008 | -0.974 | 0.330 | 0.867 |
|  | TrailA | 0.004 | 0.006 | 0.798 | 0.425 | 0.867 |
|  | TrailB | 0.004 | 0.005 | 0.821 | 0.411 | 0.867 |
| Hysterectomy * *APOE* ε4 | PairsMatch | 0.090 | 0.082 | 1.097 | 0.273 | 0.867 |
|  | ReactionTime | -0.125 | 0.078 | -1.609 | 0.108 | 0.867 |
|  | SymbolSub | 0.127 | 0.170 | 0.743 | 0.457 | 0.867 |
|  | DigitSpan | -0.400 | 0.231 | -1.729 | 0.084 | 0.867 |
|  | TrailA | 0.335 | 0.183 | 1.829 | 0.067 | 0.862 |
|  | TrailB | 0.058 | 0.178 | 0.324 | 0.746 | 0.947 |
| Age at Hysterectomy * *APOE* ε4 | PairsMatch | 0.002 | 0.003 | 0.697 | 0.486 | 0.867 |
|  | ReactionTime | -0.006 | 0.003 | -2.245 | 0.025 | 0.596 |
|  | SymbolSub | -0.003 | 0.004 | -0.605 | 0.545 | 0.867 |
|  | DigitSpan | 0.006 | 0.008 | 0.674 | 0.501 | 0.867 |
|  | TrailA | -0.009 | 0.005 | -1.800 | 0.072 | 0.862 |
|  | TrailB | -0.001 | 0.005 | -0.115 | 0.908 | 0.977 |
| N Childbirth * *APOE* ε4 | PairsMatch | -0.008 | 0.011 | -0.718 | 0.473 | 0.867 |
|  | ReactionTime | 0.039 | 0.011 | 3.589 | **3.32e-04** | **0.022** |
|  | SymbolSub | 0.031 | 0.022 | 1.436 | 0.151 | 0.867 |
|  | DigitSpan | 0.012 | 0.041 | 0.294 | 0.769 | 0.958 |
|  | TrailA | -0.022 | 0.025 | -0.886 | 0.375 | 0.867 |
|  | TrailB | -0.021 | 0.024 | -0.887 | 0.375 | 0.867 |
| N Childbirth^2^ * *APOE* ε4 | PairsMatch | 0.009 | 0.012 | 0.721 | 0.471 | 0.867 |
|  | ReactionTime | -0.046 | 0.012 | -3.831 | **1.28e-04** | **0.017** |
|  | SymbolSub | -0.032 | 0.027 | -1.182 | 0.237 | 0.867 |
|  | DigitSpan | -0.040 | 0.047 | -0.838 | 0.402 | 0.867 |
|  | TrailA | 0.015 | 0.031 | 0.477 | 0.633 | 0.867 |
|  | TrailB | 0.001 | 0.030 | 0.047 | 0.963 | 0.983 |
| Age last Birth * *APOE* ε4 | PairsMatch | 0.043 | 0.021 | 2.054 | 0.040 | 0.755 |
|  | ReactionTime | 0.000 | 0.020 | -0.002 | 0.999 | 0.999 |
|  | SymbolSub | 0.027 | 0.038 | 0.716 | 0.474 | 0.867 |
|  | DigitSpan | 0.037 | 0.065 | 0.567 | 0.571 | 0.867 |
|  | TrailA | -0.002 | 0.045 | -0.045 | 0.964 | 0.983 |
|  | TrailB | 0.042 | 0.043 | 0.962 | 0.336 | 0.867 |
| Age first Birth * *APOE* ε4 | PairsMatch | -0.005 | 0.021 | -0.234 | 0.815 | 0.966 |
|  | ReactionTime | 0.013 | 0.020 | 0.656 | 0.512 | 0.867 |
|  | SymbolSub | -0.007 | 0.039 | -0.179 | 0.858 | 0.966 |
|  | DigitSpan | -0.068 | 0.061 | -1.107 | 0.269 | 0.867 |
|  | TrailA | 0.004 | 0.047 | 0.092 | 0.927 | 0.983 |
|  | TrailB | 0.009 | 0.046 | 0.205 | 0.837 | 0.966 |
| Miscarriage & Termination * *APOE* ε4 | PairsMatch | -0.025 | 0.019 | -1.271 | 0.204 | 0.867 |
|  | ReactionTime | 0.003 | 0.018 | 0.149 | 0.881 | 0.970 |
|  | SymbolSub | -0.030 | 0.034 | -0.859 | 0.390 | 0.867 |
|  | DigitSpan | 0.055 | 0.062 | 0.887 | 0.375 | 0.867 |
|  | TrailA | -0.015 | 0.041 | -0.374 | 0.709 | 0.908 |
|  | TrailB | -0.016 | 0.039 | -0.414 | 0.679 | 0.896 |
| Stillbirth * *APOE* ε4 | PairsMatch | 0.020 | 0.019 | 1.062 | 0.288 | 0.867 |
|  | ReactionTime | 0.011 | 0.018 | 0.600 | 0.549 | 0.867 |
|  | SymbolSub | 0.021 | 0.034 | 0.618 | 0.536 | 0.867 |
|  | DigitSpan | 0.038 | 0.062 | 0.608 | 0.543 | 0.867 |
|  | TrailA | -0.033 | 0.042 | -0.783 | 0.434 | 0.867 |
|  | TrailB | -0.002 | 0.040 | -0.061 | 0.951 | 0.983 |
| Current HT use * *APOE* ε4 | PairsMatch | -0.007 | 0.010 | -0.627 | 0.531 | 0.867 |
|  | ReactionTime | 0.007 | 0.010 | 0.741 | 0.459 | 0.867 |
|  | SymbolSub | -0.018 | 0.017 | -1.067 | 0.286 | 0.867 |
|  | DigitSpan | -0.025 | 0.034 | -0.737 | 0.461 | 0.867 |
|  | TrailA | -0.014 | 0.020 | -0.711 | 0.477 | 0.867 |
|  | TrailB | -0.010 | 0.019 | -0.530 | 0.596 | 0.867 |
| Past HT use * *APOE* ε4 | PairsMatch | 0.003 | 0.006 | 0.459 | 0.646 | 0.867 |
|  | ReactionTime | 0.008 | 0.005 | 1.486 | 0.137 | 0.867 |
|  | SymbolSub | -0.022 | 0.010 | -2.275 | 0.023 | 0.596 |
|  | DigitSpan | -0.009 | 0.017 | -0.539 | 0.590 | 0.867 |
|  | TrailA | 0.003 | 0.011 | 0.236 | 0.814 | 0.966 |
|  | TrailB | -0.001 | 0.011 | -0.113 | 0.910 | 0.977 |
| Age HT initiation * *APOE* ε4 | PairsMatch | -0.004 | 0.004 | -0.908 | 0.364 | 0.867 |
|  | ReactionTime | 0.003 | 0.004 | 0.712 | 0.476 | 0.867 |
|  | SymbolSub | -0.012 | 0.008 | -1.518 | 0.129 | 0.867 |
|  | DigitSpan | -0.003 | 0.013 | -0.239 | 0.811 | 0.966 |
|  | TrailA | -0.006 | 0.010 | -0.634 | 0.526 | 0.867 |
|  | TrailB | -0.002 | 0.009 | -0.248 | 0.804 | 0.966 |
| Age HT rel. Menopause * *APOE* ε4 | PairsMatch | -0.002 | 0.004 | -0.458 | 0.647 | 0.867 |
|  | ReactionTime | 0.002 | 0.004 | 0.571 | 0.568 | 0.867 |
|  | SymbolSub | -0.004 | 0.008 | -0.467 | 0.640 | 0.867 |
|  | DigitSpan | -0.003 | 0.013 | -0.198 | 0.843 | 0.966 |
|  | TrailA | 0.007 | 0.009 | 0.809 | 0.419 | 0.867 |
|  | TrailB | 0.000 | 0.009 | -0.031 | 0.975 | 0.983 |
| Duration HT use * *APOE* ε4 | PairsMatch | -0.005 | 0.005 | -1.067 | 0.286 | 0.867 |
|  | ReactionTime | 0.004 | 0.005 | 0.796 | 0.426 | 0.867 |
|  | SymbolSub | 0.000 | 0.009 | -0.032 | 0.975 | 0.983 |
|  | DigitSpan | -0.003 | 0.016 | -0.172 | 0.864 | 0.966 |
|  | TrailA | 0.019 | 0.012 | 1.645 | 0.100 | 0.867 |
|  | TrailB | 0.022 | 0.011 | 1.944 | 0.052 | 0.761 |
| Current HC use * *APOE* ε4 | PairsMatch | 0.005 | 0.018 | 0.306 | 0.759 | 0.955 |
|  | ReactionTime | -0.008 | 0.017 | -0.454 | 0.650 | 0.867 |
|  | SymbolSub | 0.066 | 0.028 | 2.354 | 0.019 | 0.596 |
|  | DigitSpan | 0.064 | 0.054 | 1.178 | 0.239 | 0.867 |
|  | TrailA | 0.001 | 0.032 | 0.041 | 0.968 | 0.983 |
|  | TrailB | 0.048 | 0.031 | 1.546 | 0.122 | 0.867 |
| Past HC use * *APOE* ε4 | PairsMatch | 0.003 | 0.006 | 0.479 | 0.632 | 0.867 |
|  | ReactionTime | -0.001 | 0.006 | -0.180 | 0.857 | 0.966 |
|  | SymbolSub | 0.017 | 0.012 | 1.418 | 0.156 | 0.867 |
|  | DigitSpan | 0.033 | 0.021 | 1.573 | 0.116 | 0.867 |
|  | TrailA | 0.019 | 0.014 | 1.346 | 0.178 | 0.867 |
|  | TrailB | 0.018 | 0.013 | 1.305 | 0.192 | 0.867 |
| Age HC initiation * *APOE* ε4 | PairsMatch | 0.003 | 0.003 | 1.056 | 0.291 | 0.867 |
|  | ReactionTime | -0.001 | 0.003 | -0.218 | 0.827 | 0.966 |
|  | SymbolSub | -0.003 | 0.006 | -0.518 | 0.605 | 0.867 |
|  | DigitSpan | 0.008 | 0.010 | 0.791 | 0.429 | 0.867 |
|  | TrailA | -0.006 | 0.007 | -0.799 | 0.424 | 0.867 |
|  | TrailB | -0.006 | 0.007 | -0.804 | 0.421 | 0.867 |
| Duration HC use * *APOE* ε4 | PairsMatch | 0.004 | 0.003 | 1.186 | 0.236 | 0.867 |
|  | ReactionTime | -0.002 | 0.003 | -0.755 | 0.450 | 0.867 |
|  | SymbolSub | -0.004 | 0.006 | -0.667 | 0.505 | 0.867 |
|  | DigitSpan | 0.004 | 0.010 | 0.398 | 0.691 | 0.897 |
|  | TrailA | -0.004 | 0.007 | -0.504 | 0.614 | 0.867 |
|  | TrailB | -0.004 | 0.007 | -0.507 | 0.612 | 0.867 |

Abbreviation: APOE = Apolipoprotein. PairMatch = Pair Matching Test. ReactionTime = Reaction Time Test. SymbolSub = Symbol Substitution Test. DigitSpan = Digit Span Test. Trail A & B = Trail Making Test A & B. N = Number. HT = Hormone Therapy. HC = Hormonal Contraceptive. S.E. = Standard Error. FDR = False Discovery Rate. Significant results are highlighted in bold.

1.	Foster HME, Celis-Morales CA, Nicholl BI, Petermann-Rocha F, Pell JP, Gill JMR *et al.* The effect of socioeconomic deprivation on the association between an extended measurement of unhealthy lifestyle factors and health outcomes: a prospective analysis of the UK Biobank cohort. *Lancet Public Health* 2018; **3**(12)**:** e576-e585.
